# Understanding care-seeking of pregnant women from underserved groups: A systematic review and meta-ethnography

**DOI:** 10.1101/2025.07.24.25332124

**Authors:** Tisha Dasgupta, Hannah Rayment-Jones, Gillian Horgan, Yesmin Begum, Michelle Peter, Sergio A. Silverio, Laura A Magee

## Abstract

**Background:** Delayed or reduced antenatal care use by pregnant women may result in poorer outcomes. ‘Candidacy’ is a synthetic framework which outlines how people’s eligibility for healthcare is jointly negotiated. This meta-ethnography aimed to identify – through the lens of candidacy – factors affecting experiences of care-seeking during pregnancy by women from underserved communities in high-income countries (HICs).

**Methods:** Six electronic databases were systematically searched, extracting papers published from January 2018 to January 2023, updated to May 2025, and having relevant qualitative data from marginalised and underserved groups in HICs. Methodological quality of included papers was assessed using the Critical Appraisal Skills Programme. Meta-ethnography was used for analytic synthesis and findings were mapped to the Candidacy Framework.

**Results:** Studies (N=51), with data from 1,347 women across 14 HICs were included. A total of 12 sub-themes across five themes were identified: (1) Autonomy, dignity, and personhood; (2) Informed choice and decision-making; (3) Trust *in* and relationship *with* healthcare professionals; (4) Differences in healthcare systems and cultures; and (5) Systemic barriers. Candidacy constructs to which themes were mapped were predominantly joint- (navigation of health system), health system- (permeability of services), and individual-level (appearances at health services). Mapping to Candidacy Framework was partial for seven sub-themes, particularly for individuals with a personal or family history of migration. The meta-ethnography allowed for the theory: ‘Respect, informed choice, and trust enhances candidacy whilst differences in healthcare systems, culture, and systemic barriers have the propensity to diminish it’.

**Conclusion:** Improvements in antenatal care utilisation must focus on the joint (service-user and -provider) nature of responsibility for care-seeking, through co-production. We suggest two additional Candidacy Framework constructs: ‘intercultural dissonance’ and ‘hostile bureaucracy’, which reflect the multi-generational impact of migration on healthcare utilisation and the intersection of healthcare utilisation with a hostile and bureaucratic environment.

**Funding:** ESRC Doctoral training fellowship (ES/P000703/1)

**Registration:** This review was registered with PROSPERO [CRD42023389306].

**Research in Context:** *Evidence before the study:* Delayed or reduced utilisation of healthcare during the perinatal period can be detrimental for both the mother and baby. Women from marginalised and underserved communities face increased barriers to seeking and engaging with care during pregnancy, which were likely further exacerbated, disproportionately, by global changes in maternity care services during the COVID-19 pandemic. A search of six electronic databases was conducted for eligible qualitative research studies published between 2018-2025 in high-income countries (HICs), to investigate factors affecting experiences of care-seeking during pregnancy, by women and birthing people from underserved communities. The Candidacy framework was used as a theoretical lens to interrogate the data, to understand the dynamic process by which people’s eligibility for healthcare is jointly negotiated between themselves and the health system.

*Added value of this study:* Drawing from 51 published studies with data from 1,347women in 14 countries, this systematic review and meta-ethnography led to the development of a theory: *Respect, informed choice, and trust enhances candidacy whilst differences in healthcare systems, culture, and systemic barriers have the propensity to diminish it.* We add to the existing literature by providing an in-depth analysis of barriers and facilitators of care-seeking behaviour amongst a population with high levels of social complexity. Using the lens of Candidacy, we observed a dominance of connections across joint- and health system-level factors as compared to individual-level ones, emphasising joint responsibility for positive experiences of maternity care-seeking. Furthermore, we propose two new constructs of ‘intercultural dissonance’ and ‘hostile bureaucracy’ to be added to the Candidacy framework, as emerging of particular relevance to migrants, reflecting intergenerational relationship changes and hostile immigration policies faced by these individuals.

*Implications of all the available evidence:* The present synthesis emphasises the need for policy and practice improvements in maternity care utilisation, which focus on the joint (service-user and -provider) nature of responsibility for care-seeking, through co-production. In particular, events of the last decade have emphasised the underserved nature of migrants, refugees, and asylum seekers; a population which has grown exponentially in the recent past due to various humanitarian crises, and are in need of additional support in maternity care services in HICs.

## Introduction

Routine antenatal care is a globally recommended public health service enabling healthcare professionals (HCPs) to provide essential information, counselling, maternal and fetal assessments, and encourage use of maternity services.^1,2^ Delayed or reduced antenatal care use, in both high- (HICs) and low-/middle-income countries (LMICs), is linked to adverse pregnancy outcomes, including stillbirth,^3^ and neonatal morbidity.^4^

Research on maternity care-seeking has largely focused on LMICs, where barriers are often financial, geographic, or linked to knowledge gaps beliefs about the importance of maternity care.^5^ In contrast, many HICs offer free healthcare at point of access, yet barriers remain. Even with structurally accessible services, uptake remains low in certain communities, including those of lower socio-economic status (SES), minority ethnic groups, sexual minorities, and people living with disabilities.^6–8^

A recent meta-synthesis of qualitative studies in HICs highlighted multiple barriers (e.g., socio-demographic disadvantage, system navigation, lack of tailored care, frequent carer changes) and facilitators (e.g., positive pregnancy attitudes, good HCP interactions, social support).^9^ Furthermore, the pandemic introduced additional barriers to care-seeking (i.e., social isolation, personal infection risk,^10^ poorer mental well-being,^11^ continuing restrictions for perinatal populations after lockdowns,^12^ and navigating healthcare service reconfigurations^13,14^), with experiences of care being reported more negatively with poorer mental health outcomespoorer.^11–18^

‘Candidacy’ refers to people’s eligibility for accessing healthcare. It was developed to explain unequal access to healthcare, despite universal health coverage, and to go beyond simple measurement of health utilisation, particularly by marginalised groups.^19^ The theoretical framework of ‘candidacy’ refers to healthcare access as negotiated jointly between service-user and healthcare system. It describes a dynamic process, subject to external influences, from people and their social context, as well as available resources and service structure.^19^ There are seven constructs of: identification, navigation, permeability of services, appearances at health services, adjudication, offers and resistance, and local production of candidacy.^19^ As used previously in healthcare research, this framework lends itself well to understanding the latent factors influencing care-seeking amongst marginalised groups, for which it was first developed.^20–25^

The aim of this systematic review and meta-ethnography was to synthesise qualitative evidence from HICs, to identify – through the lens of candidacy – factors affecting experiences of care-seeking during pregnancy, by women and birthing people from underserved communities. We expand on previous work by focusing solely on underserved groups known to face additional barriers to care access, utilisation, and engagement.

## Methods

This review was registered with PROSPERO [CRD42023389306] and adheres to the PRISMA 2020 statement (**Table S1**).^26^

### Inclusion and exclusion criteria

The PEO (Population, Exposure, Outcome) framework was used to formulate the search strategy as per the research aim (**Table S2**). Study designs included: descriptive, exploratory, and interpretive qualitative studies; ethnographies; and observational or mixed-methods studies (including surveys with open-ended questions) where qualitative data had been formally analysed and presented.^24^ Studies were published between Jan 2018-23, updated to May 2025, and only considered if published in English-language.

Studies of postnatal care were excluded due to its variation between countries, and its fragmented nature, often spanning services in primary through to quaternary care settings.

### Search strategy and selection

Electronic databases of SCOPUS, MEDLINE, EMBASE, CINAHL, Global Health, PsychINFO, and MIDRIS were systematically searched for articles published between 1 January 2018 and 1 January 2023, updated to May 2025. Dates were selected to align with the wider study’s quantitative data analysis time-frame.^27^ For details of the search terms and keywords used, see **Table S2**.

Duplicate references were removed using Mendeley reference manager software, and citations were uploaded to Rayyan,^28^ a web-based tool for conducting systematic reviews. At least two members of the study team (TD, HRJ, GH, SAS, LAM) independently screened each record, by title and abstract, followed by full-text review. Regular discussions were held to resolve by consensus any disagreements in screening decisions.

### Data extraction

Data extraction was randomly allocated to one of two reviewers (TD, GH), with 20% of included studies extracted independently by both reviewers to check between-reviewer reliability. A bespoke Microsoft Excel spreadsheet was used to abstract study characteristics (i.e., title, reference, publication year, study setting, aim, participant inclusion criteria, intersectional approach, data collection and analytic methodologies), and any impact of the pandemic on care-seeking. Regular discussions were organised to discuss any disagreements, and to collaborate on the creation of a consolidated set of themes with consistent labels.

### Quality assessment

The Critical Appraisal Skills Programme (CASP) was used to assess the quality of included studies^29^ across ten items: clearly-stated objective, appropriateness of using qualitative study design, justification of research design, recruitment strategy, data collection method, author reflexivity, ethical considerations, data analysis method, clear findings, and value of the findings. CASP does not assign a score, but for ease of interpretation, we assigned points to answers for each checklist item: 0 points for ‘No’, 1 for ‘Cannot tell’, and 2 for ‘Yes’.

### Data synthesis

Meta-ethnography^30^ was employed for analytic data synthesis, which is a particularly useful approach when addressing complex questions, as it enables comparison between and across published studies, and creates higher-order themes which can be newly-interpreted, based on the wealth of integrated data.^30,31^ Syntheses can be reciprocal (studies are similar to each other and shared themes across the studies are summarised), refutational (studies refute each other and themes are juxtaposed against each other), or ‘line of argument’ (studies interpret the same phenomenon but from different aspects, the synthesis creating a whole greater than the sum of its individual parts).^31^ Typically, there are four main steps, as employed by other researchers^32,33^, outlined below in **Figure 1**:^31–33^

**Figure 1.**
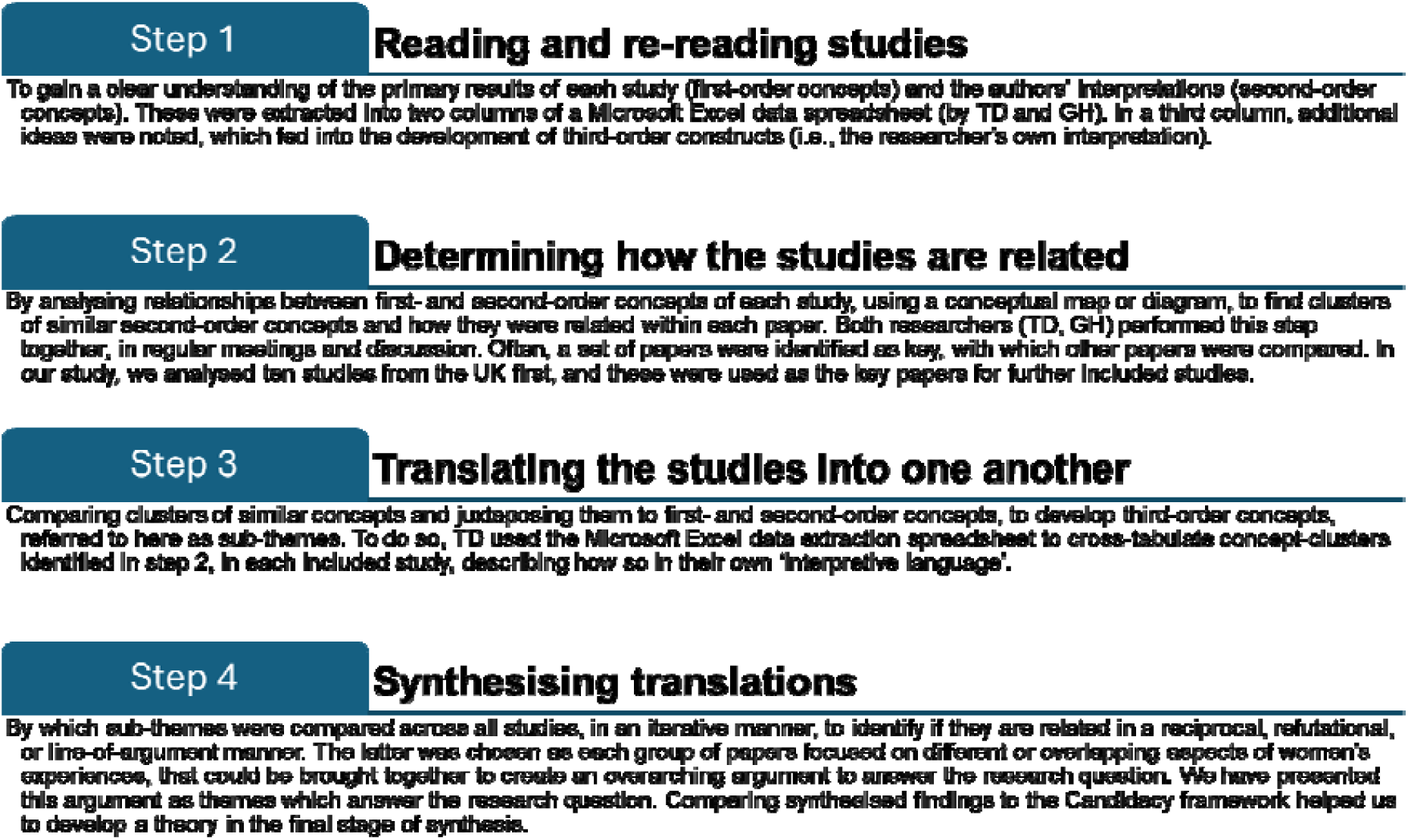
Steps of meta-ethnography.

Intersectional approaches in included studies were considered to compare participant groups. Synthesised themes and sub-themes were mapped to one or more of the seven components of the Candidacy Framework,^19^ as in **Table 1** below, with the weight of each theme contributing to candidacy was calculated.

**Table 1:**
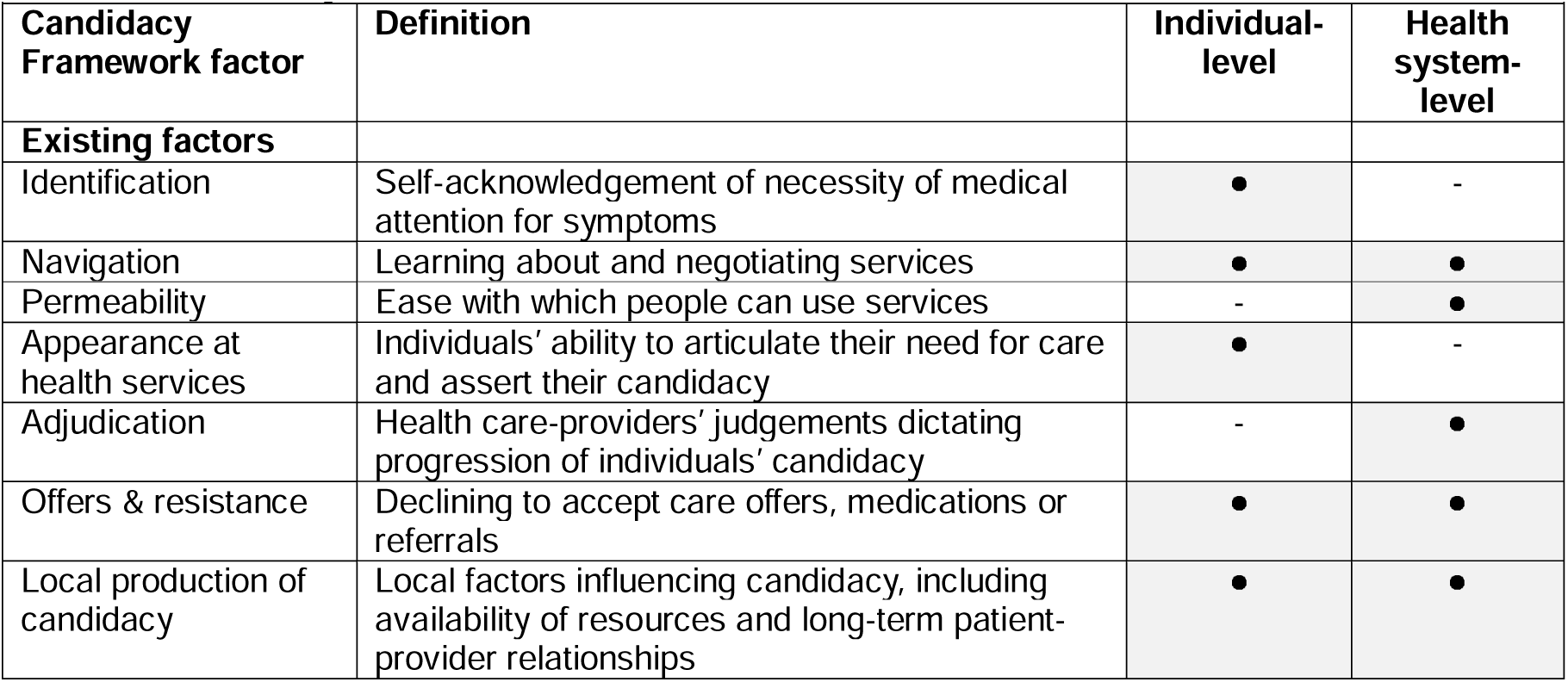
Candidacy framework constructs.

## Results

### Search and selection

Of 3,098 records identified, 2,493 underwent title and abstract screening, 68 underwent full-text review, and 45 were included.^34–78^ (see **Figure S1**). An updated search to May 2025, identified six additional records for analysis.^79–84^

### Description of included studies

The 51 included studies provided data from 1,347 service-users. Studies were published between 2018-2025, from 14 countries, most commonly the USA (n=13 studies)^35–37,45,51,52,56,60,63,66,67,79^ and the UK (n=13),^57,69–78,83,84^ followed in frequency by Australia (n=5),^42,48,55,65,81^ Norway,^46,49,68^ Denmark,^44,61,62^ Sweden,^34,47,50^ Switzerland,^40,41^ Netherlands,^39,64^ New Zealand,^43,80^ Canada,^38^ Germany,^59^ Israel,^54^ Russia,^82^ and Saudi Arabia.^58^ The most common data collection method was in-depth interviews (n=34),^34,38–40,42,44,47–50,52–59,61–63,66–68,70,74,75,77–82,84^ followed by focus groups (n=13),^35–37,41,43,45,46,51,60,64,69,71,73^ surveys with open-ended questions,^65,72,76,83^ and ethnographic observations.^69^ Some studies used multiple methods (n=6).^35,43,45,46,69,73^ Most studies utilised thematic analyses (n=31);^36,37,40,41,43,46,51,53–58,60,62,64,65,67–70,72,73,76–81,83,84^ others used framework analyses (n=5),^39,63,71,74,75^ content analyses (n=5),^34,47,50,59,66^ grounded theory analysis (n=4),^35,38,45,52^ interpretative phenomenological analysis,^48,82^ systematic text condensation,^44,49^ qualitative comparative analysis,^42,56^ or interpretive description analysis.^67^ Two studies^50,66^ evaluated the impact of the pandemic on care-seeking experiences.

Social risk factors included: being migrants, refugees, or asylum-seekers (n=18);^34,35,41,44–47,49,50,55,59,64,66–69,73,74,78 38,42,63–65,73–75,78,81^ being racial, ethnic or religious minorities (n=9);^36–38,43,51,53,54,71,80 55,59,60,64,65,68,81,83,85–87^ having medical complexity (n=9);^35,37,39,47,51,55,60–62^ having low SES (n=8);^70–72,75,78–81^ not being able to speak the local language (n=7);^40,50,53,56,57,59,65,84^ having previous interaction with social services or child protection services (n=4);^61,62,75,83^ having substance abuse issues (n=4);^52,61,62,83,55,64,65,86^ having learning, intellectual, or physical disability or impairment (n=3);^76,80,83^ being a victim of domestic abuse or intimate partner violence (n=3);^42,78,83^ being a young mother (n=3);^78,80,83^ living in a rural setting (n=3);^51,81,82,64,65,78,86^ having missed or delayed antenatal care (n=2);^58,63^ experiencing homelessness (n=2);^77,83^ or having transgender pregnancy (n=1).^80^ For further details, see **Table S3**.

### Quality Assessment

Study quality was moderate-to-high (**Table S4**). Of a possible score of 20, all studies scored ≥14, as follows: ^60^14/20 (n=2),^38,65^ 15/20 (n=4),^34,53,74,83^ 16/20 (n=4),^52,60,68,72^ 17/20 (n=15),^35,43,46–48,56,58,59,62,64,66,73,75,77,78^ 18/20 (n=13),^40,41,44,49,51,54,55,63,70,71,76,82,84^ 19/20 (n=5),^36,42,45,57,79,80^ and 20/20 (n=7).^37,39,50,61,67,69,81^ Those highest-scoring studies which did not reach 20/20 often fell short by missing consideration of the relationship between researchers and participants and associated ethical issues.

### Analytic Synthesis and Findings

Figure 2 depicts the theory developed: ‘Respect, informed choice, and trust enhances candidacy whilst differences in healthcare systems, culture, and systemic barriers have the propensity to diminish it’. The 12 sub-themes were grouped into five main themes: (1) Autonomy, dignity, and personhood; (2) Informed choice and decision-making; (3) Trust in and relationship with HCP; (4) Differences in healthcare systems and cultures; and (5) Systemic factors. Excerpts of text from individual studies are presented in **Table 2** to support the synthesised findings (direct participant quotations are in italics).

**Figure 2.**
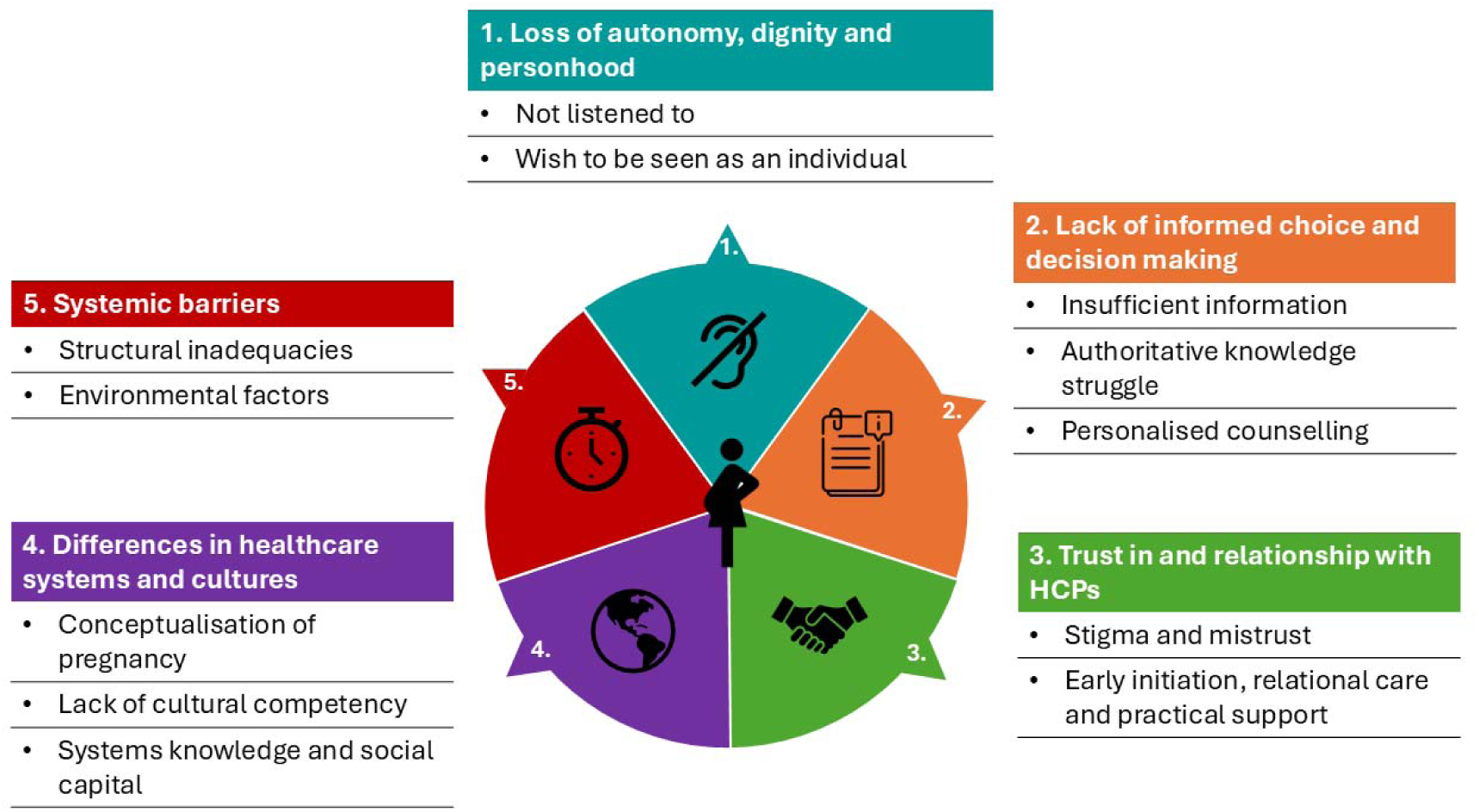
Findings of factors affecting women’s experience of care-seeking.

**Table 2.**
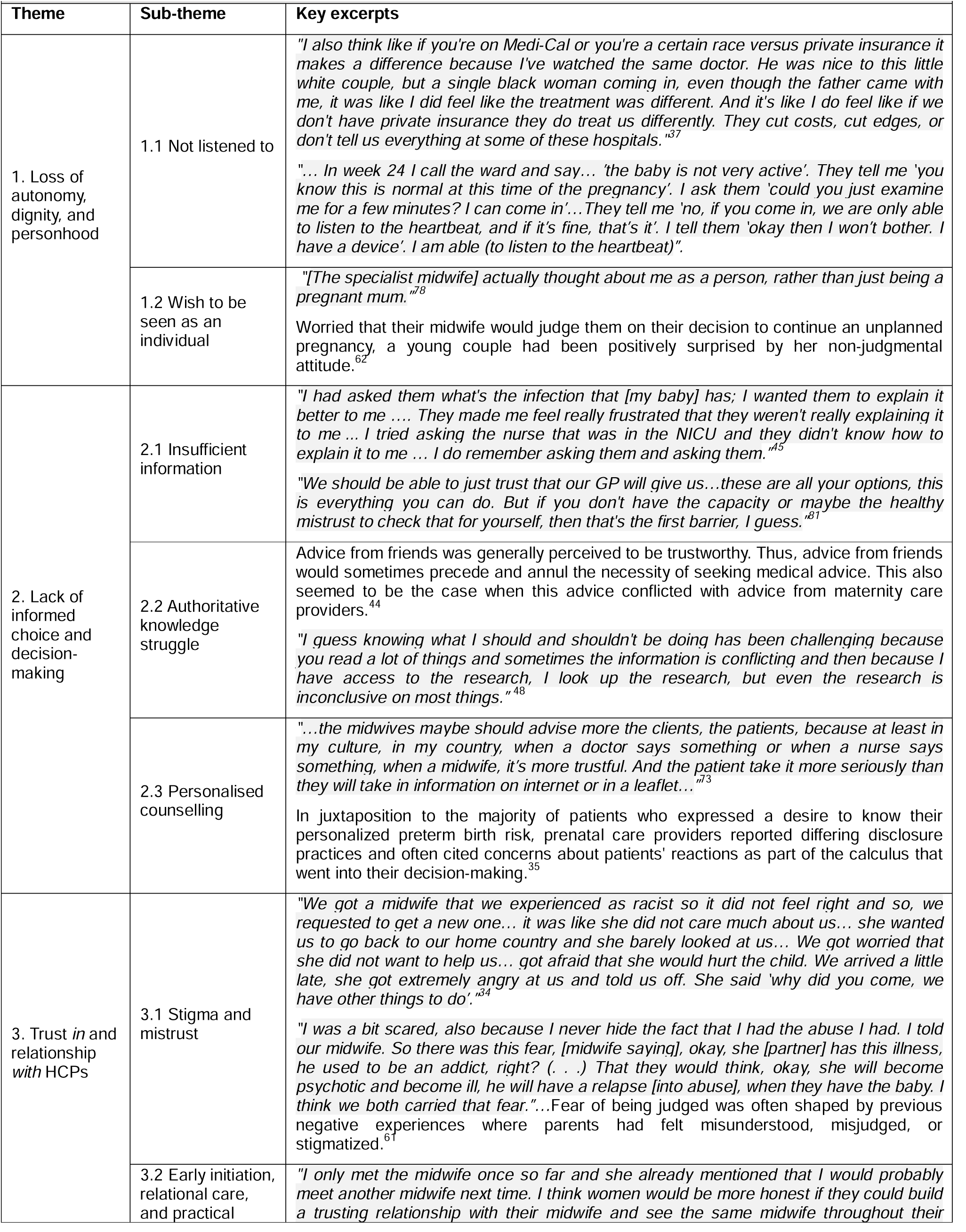
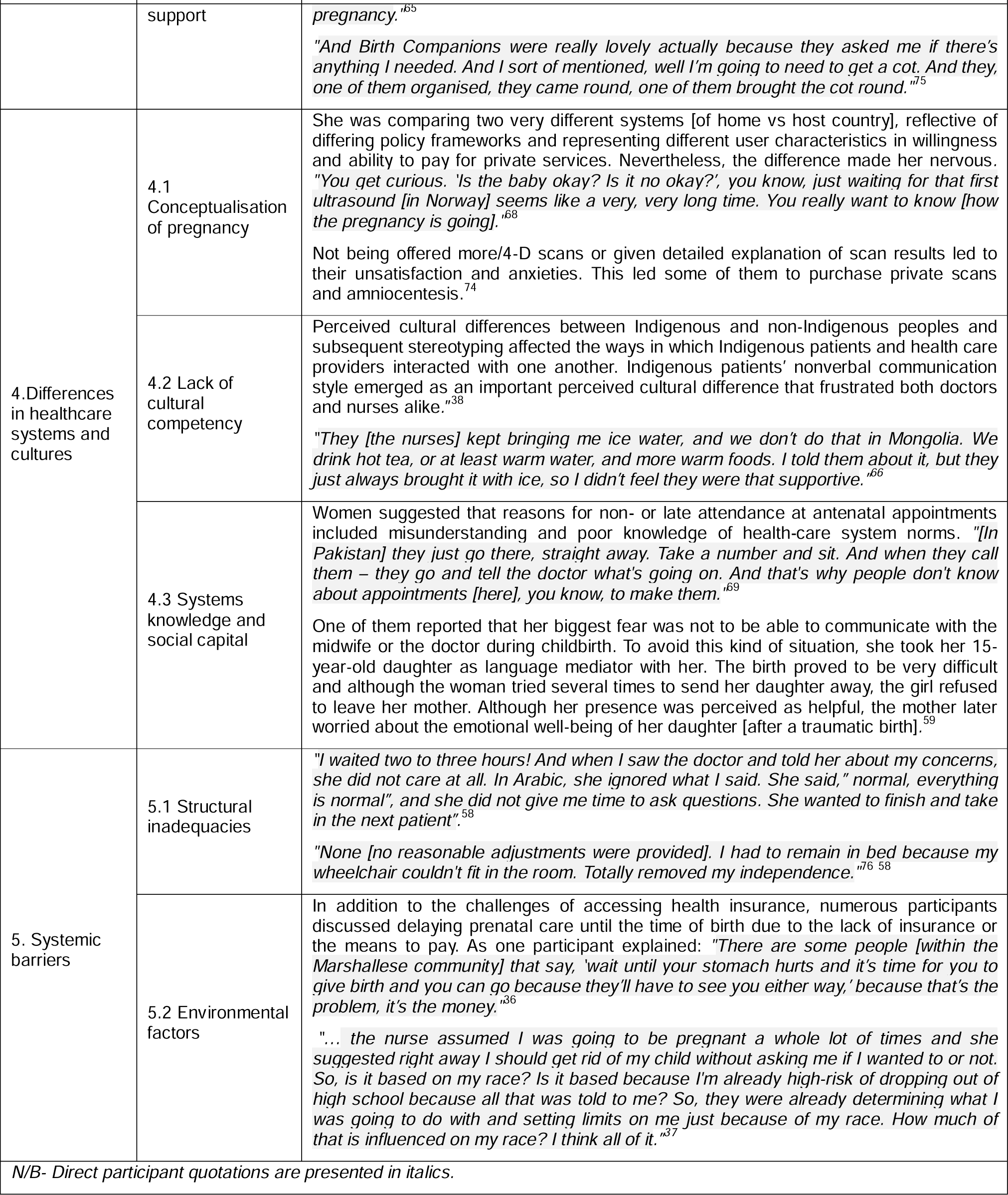
Key quotations to support thematic findings.

| Table 2. Key quotations to support thematic findings |  |  |
| --- | --- | --- |
| Theme | Sub-theme | Key excerpts |
| 1. Loss of autonomy, dignity, and personhood | 1.1 Not listened to | "I also think like if you're on Medi-Cal or you're a certain race versus private insurance it makes a difference because I've watched the same doctor. He was nice to this little white couple, but a single black woman coming in, even though the father came with me, it was like I did feel like the treatment was different. And it's like I do feel like if we don't have private insurance they do treat us differently. They cut costs, cut edges, or don't tell us everything at some of these hospitals."<sup>37</sup><br>"... In week 24 I call the ward and say... 'the baby is not very active'. They tell me 'you know this is normal at this time of the pregnancy'. I ask them 'could you just examine me for a few minutes? I can come in'... They tell me 'no, if you come in, we are only able to listen to the heartbeat, and if it's fine, that's it'. I tell them 'okay then I won't bother. I have a device'. I am able (to listen to the heartbeat)". |
|  | 1.2 Wish to be seen as an individual | "[The specialist midwife] actually thought about me as a person, rather than just being a pregnant mum."<sup>78</sup><br>Worried that their midwife would judge them on their decision to continue an unplanned pregnancy, a young couple had been positively surprised by her non-judgmental attitude.<sup>62</sup> |
| 2. Lack of informed choice and decision-making | 2.1 Insufficient information | "I had asked them what's the infection that [my baby] has; I wanted them to explain it better to me .... They made me feel really frustrated that they weren't really explaining it to me ... I tried asking the nurse that was in the NICU and they didn't know how to explain it to me ... I do remember asking them and asking them."<sup>45</sup><br>"We should be able to just trust that our GP will give us...these are all your options, this is everything you can do. But if you don't have the capacity or maybe the healthy mistrust to check that for yourself, then that's the first barrier, I guess."<sup>81</sup> |
|  | 2.2 Authoritative knowledge struggle | Advice from friends was generally perceived to be trustworthy. Thus, advice from friends would sometimes precede and annul the necessity of seeking medical advice. This also seemed to be the case when this advice conflicted with advice from maternity care providers.<sup>44</sup><br>"I guess knowing what I should and shouldn't be doing has been challenging because you read a lot of things and sometimes the information is conflicting and then because I have access to the research, I look up the research, but even the research is inconclusive on most things."<sup>48</sup> |
|  | 2.3 Personalised counselling | "...the midwives maybe should advise more the clients, the patients, because at least in my culture, in my country, when a doctor says something or when a nurse says something, when a midwife, it's more trustful. And the patient take it more seriously than they will take in information on internet or in a leaflet..."<sup>73</sup><br>In juxtaposition to the majority of patients who expressed a desire to know their personalized preterm birth risk, prenatal care providers reported differing disclosure practices and often cited concerns about patients' reactions as part of the calculus that went into their decision-making.<sup>35</sup> |
| 3. Trust in and relationship with HCPs | 3.1 Stigma and mistrust | "We got a midwife that we experienced as racist so it did not feel right and so, we requested to get a new one... it was like she did not care much about us... she wanted us to go back to our home country and she barely looked at us... We got worried that she did not want to help us... got afraid that she would hurt the child. We arrived a little late, she got extremely angry at us and told us off. She said 'why did you come, we have other things to do'."<sup>34</sup><br>"I was a bit scared, also because I never hide the fact that I had the abuse I had. I told our midwife. So there was this fear, [midwife saying], okay, she [partner] has this illness, he used to be an addict, right? (. . .) That they would think, okay, she will become psychotic and become ill, he will have a relapse [into abuse], when they have the baby. I think we both carried that fear..." Fear of being judged was often shaped by previous negative experiences where parents had felt misunderstood, misjudged, or stigmatized.<sup>61</sup> |
|  | 3.2 Early initiation, relational care, and practical | "I only met the midwife once so far and she already mentioned that I would probably meet another midwife next time. I think women would be more honest if they could build a trusting relationship with their midwife and see the same midwife throughout their |
| Theme | Sub-theme | Key excerpts |
|  | support | *pregnancy.*<sup>65</sup><br>*"And Birth Companions were really lovely actually because they asked me if there's anything I needed. And I sort of mentioned, well I'm going to need to get a cot. And they, one of them organised, they came round, one of them brought the cot round."*<sup>75</sup> |
| 4. Differences in healthcare systems and cultures | 4.1 Conceptualisation of pregnancy | She was comparing two very different systems [of home vs host country], reflective of differing policy frameworks and representing different user characteristics in willingness and ability to pay for private services. Nevertheless, the difference made her nervous. *"You get curious. 'Is the baby okay? Is it no okay?'; you know, just waiting for that first ultrasound [in Norway] seems like a very, very long time. You really want to know [how the pregnancy is going]."*<sup>68</sup><br>Not being offered more/4-D scans or given detailed explanation of scan results led to their unsatisfaction and anxieties. This led some of them to purchase private scans and amniocentesis.<sup>74</sup> |
|  | 4.2 Lack of cultural competency | Perceived cultural differences between Indigenous and non-Indigenous peoples and subsequent stereotyping affected the ways in which Indigenous patients and health care providers interacted with one another. Indigenous patients' nonverbal communication style emerged as an important perceived cultural difference that frustrated both doctors and nurses alike.<sup>38</sup><br>*"They [the nurses] kept bringing me ice water, and we don't do that in Mongolia. We drink hot tea, or at least warm water, and more warm foods. I told them about it, but they just always brought it with ice, so I didn't feel they were that supportive."*<sup>66</sup> |
|  | 4.3 Systems knowledge and social capital | Women suggested that reasons for non- or late attendance at antenatal appointments included misunderstanding and poor knowledge of health-care system norms. *"[In Pakistan] they just go there, straight away. Take a number and sit. And when they call them – they go and tell the doctor what's going on. And that's why people don't know about appointments [here], you know, to make them."*<sup>69</sup><br>One of them reported that her biggest fear was not to be able to communicate with the midwife or the doctor during childbirth. To avoid this kind of situation, she took her 15-year-old daughter as language mediator with her. The birth proved to be very difficult and although the woman tried several times to send her daughter away, the girl refused to leave her mother. Although her presence was perceived as helpful, the mother later worried about the emotional well-being of her daughter [after a traumatic birth].<sup>59</sup> |
| 5. Systemic barriers | 5.1 Structural inadequacies | *"I waited two to three hours! And when I saw the doctor and told her about my concerns, she did not care at all. In Arabic, she ignored what I said. She said, "normal, everything is normal", and she did not give me time to ask questions. She wanted to finish and take in the next patient"*<sup>58</sup><br>*"None [no reasonable adjustments were provided]. I had to remain in bed because my wheelchair couldn't fit in the room. Totally removed my independence."*<sup>76 58</sup> |
|  | 5.2 Environmental factors | In addition to the challenges of accessing health insurance, numerous participants discussed delaying prenatal care until the time of birth due to the lack of insurance or the means to pay. As one participant explained: *"There are some people [within the Marshallese community] that say, 'wait until your stomach hurts and it's time for you to give birth and you can go because they'll have to see you either way,' because that's the problem, it's the money."*<sup>36</sup><br>*"... the nurse assumed I was going to be pregnant a whole lot of times and she suggested right away I should get rid of my child without asking me if I wanted to or not. So, is it based on my race? Is it based because I'm already high-risk of dropping out of high school because all that was told to me? So, they were already determining what I was going to do with and setting limits on me just because of my race. How much of that is influenced on my race? I think all of it."*<sup>37</sup> |
| N/B- Direct participant quotations are presented in italics. |  |  |

#### 1. Theme 1: Loss of autonomy, dignity and personhood

This theme was identified in 16 studies,^34,37,41,44,45,54,58,62,66,70,75–78^ and had two sub-themes.

##### 1.1 Not listened to

Participants expressed they were not listened to, their concerns were dismissed, or they were made to feel unintelligent and judged for asking questions.^37^ Some attributed this treatment to personal characteristics, such as ethnicity. This led women to hesitate to ask further questions, attending appointments unless absolutely necessary, or engaging with maternity care overall.^77^ Some were unaware of their rights and the level of care to expect and request. This made women accept poor quality-of-care and discriminatory practices as part of standard maternity care.^54^

##### 1.2 Wish to be seen as an individual

Women wished to be respected and treated as individuals. They valued when effort was made to understand their background and life beyond pregnancy;^78^ this often had a protective effect on care-seeking and engagement and built capacity for positive parenting and health.^75^

#### 2. Theme 2: Lack of informed choice and decision-making

This theme was identified in 25 studies,^35,36,39,42,43,45,47–50,55–58,60,68,69,71–73,75,78^ and had three sub-themes.

##### 2.1 Insufficient information

Women felt information was inadequate, and lacked justification for recommendations, which left them wanting more control over their care.^55^ Some studies found HCPs’ own implicit bias and their perceptions of patients influenced the information they provided, so they offered only the information they deemed would be relevant for the patient.^59^ Women felt it fell to them to seek-out information (via friends and family, or online sources, often unofficial^74^), and make decisions about which recommendations to follow, although they felt those decisions were seldom fully-informed.^39^

##### 2.2 Authoritative knowledge struggle

Women often faced balancing information from various sources.^69^ This included differences in care between their home countries and their current healthcare system, between friends/family and HCPs, between care-providers, or between protocols in different hospitals.^57,69^

##### 2.3 Personalised counselling

Women emphasised the value of personalised counselling by their HCP, peer support from their communities, and HCPs having the right tools to support women and families, such as knowledge of cultural practices.^42,73^

#### 3. Theme 3: Trust *in* and relationship *with* HCPs

This theme was identified in 25 studies,^34,37–40,47,49–52,57,61,62,65,67,70,75–78^ and had two sub-themes.

##### 3.1 Stigma and mistrust

The underserved populations studied were often already anxious about being pregnant, so trust played a particularly important role in determining if they attended appointments, disclosed their circumstances, or participated in maternity care.^57^ Many had established mistrust in HCPs and institutions in general, due to prior negative interactions with social care, immigration, or law enforcement.^62,77^ Some feared being reported and their child being removed to services, and so they did not engage honestly with maternity HCPs.^77^ Women reported feeling unfit as mothers, and stigmatised when honest about social risk factors (e.g., prior drug use or homelessness).^52,67^

##### 3.2 Early initiation, relational care, and practical support

When maternity care was initiated early and there was relational care, this built trusting relationships with HCPs and facilitated open discussions.^65,77^ Women with mental health issues felt more likely to fully disclose during psychosocial assessments, and those with disabilities did not have to reiterate their accessibility requirements at every appointment.^65,77^ Practical support (e.g., with baby food, blankets, or pushchairs), or emotional support when attending social care appointments helped women embrace new motherhood.^47,67^ When they were supported in such ways, it enabled women to make long-lasting changes and prevent relapse to pre-pregnancy habits such as substance abuse.

#### 4. Theme 4: Differences in healthcare systems and cultures

This theme was identified in 21 studies,^36–38,41,44,46,47,51,55–57,59,60,68–72,74^ and had three sub-themes.

##### 4.1 Conceptualisation of pregnancy

Studies emphasised how pregnancy is conceptualised differently by setting. In some countries, antenatal care was described as highly-medicalised, with multiple appointments and ultrasound scans. In other settings, there may be only two or three contacts throughout pregnancy, even though official guidelines and recommendations may suggest more.^41,59^ Such differences often concerned mothers who had migrated from one country to another and altered their health literacy and ability to risk-assess their pregnancies. Some women did take on board new opportunities; when given the choice and relevant information, women from minority ethnic communities in the UK expressed a desire to have more home births.^71^

##### 4.2 Lack of cultural competency

Differences in social norms around pregnancy, information shared, standard practice, role of the birth partner or other family members, and religious beliefs, greatly-influenced women’s views of the acceptability of care offered, or even the decision to attend appointments.^38,46,55^ A lack of cultural understanding and respect by HCPs may have led them to perceive women’s behaviour negatively.

##### 4.3 Systems knowledge and social capital

Migrant women had trouble understanding how to access or use maternity care services in their host country, including when and how to make appointments.^36,46^ Many such women lacked social capital, described as playing a protective role, particularly postnatally. Often, they lacked support from wider familial networks during maternity care, and in life generally, to interpret for them if they did not speak the local language.^68^

#### 5. Theme 5: Systemic barriers

This theme was identified in 24 studies,^36–38,41,43,46,47,50,51,54,57–60,63,65,70,74–76^ and had two sub-themes.

##### 5.1 Structural inadequacies

Lack of flexibility in scheduling appointments, long wait-times in hospital, and rushed appointments with HCPs, posed barriers to engagement with maternity care.^43,60,70^ Studies reported poor communication between women and HCPs, due to a lack of interpreters or availability of healthcare information in other languages. Often, women resorted to methods such as Google Translate, which is not reliable for translating medical terminology, jargon, or medications.^74^ For those with physical disabilities and accessibility needs, lack of relevant provision left some women feeling that they had lost their dignity.^76^ Staff were reported as unaware of service users’ accessibility requirements (having not read their file beforehand), or unaccommodating.

##### 5.2 Environmental factors

Social, economic, political, and religious aspects played roles in how women from underserved groups were treated in hospital.^54^ Societal prejudices and systemic discriminatory practices were reported to permeate personal care interactions.^47^ In systems where care is not free-to-access at the point-of-contact (such as in the USA), even with certain health insurance plans, financial constraints deterred women from seeking care until absolutely necessary.^36^

### Contribution to the Candidacy Framework

The 12 sub-themes of this meta-ethnography mapped onto all seven components of the Candidacy Framework, with two key observations: First, most sub-themes aligned with ‘navigation’ (n=9) and ‘permeability of services’ (n=6), which are joint and health system-level influences. Fewer connections were observed with other constructs: ‘adjudication’ (n=6), ‘local production of candidacy’ (n=3), ‘offers and resistance’ (n=2), ‘appearances at health services’ (n=5), and ‘identification’ (n=3). Second, seven sub-themes only partially mapped to existing constructs: ‘authoritative knowledge struggle’, ‘stigma and mistrust’, ‘conceptualisation of pregnancy’, ‘lack of cultural competency’, ‘systems knowledge and social capital’, ‘structural inadequacies’, and ‘environmental factors’. This was especially true for those with a migrant background, suggesting the need for two additional constructs: *intercultural dissonance* (individual-level) and *hostile bureaucracy* (health system-level).

*Intercultural dissonance* encompasses additional barriers faced by those who are not native-born and experience a distinct difference in social norms and culture, medical and social knowledge and expectations, and language. Here, intergenerational relationships are altered by migration; for example, children (but not their parents) often speak (or speak more proficiently) the host country’s language, and are more familiar with the system, by virtue of having grown up there from a young age. As such, children take on more active roles in their parents’ healthcare decisions, such as acting as unofficial interpreters at care appointments, which may affect their parents’ ‘appearances at health services’ and ‘offers and resistance’ to care, as well as expose them to uncomfortable and potentially traumatic conversations and experiences.

*Hostile bureaucracy* sees migrant women often subject to discriminatory policies and precarious administrative practices in the host-country as compared to their home-country.^85^ These hostile, discriminatory immigration policies exist in most HICs, such as: restrictions on health coverage, welfare support, and right to rental properties; high visa application costs; and limits on qualifying employment. These policies, alongside negative societal attitude towards migrants and refugees, pose further barriers to integration into the host country, establishing a thriving life there, and accessing and engaging with healthcare. ‘Local production of candidacy’ is particularly diminished by these policies for migrant and refugee women.

Figure 3 shows a visual representation of the thematic contribution of our sub-themes to the original seven and extended 7+2 components of the Candidacy framework respectively. For further details of the mapping process and candidacy framework components, see **Table S5** and Table **S6** respectively.

**Figure 3.**
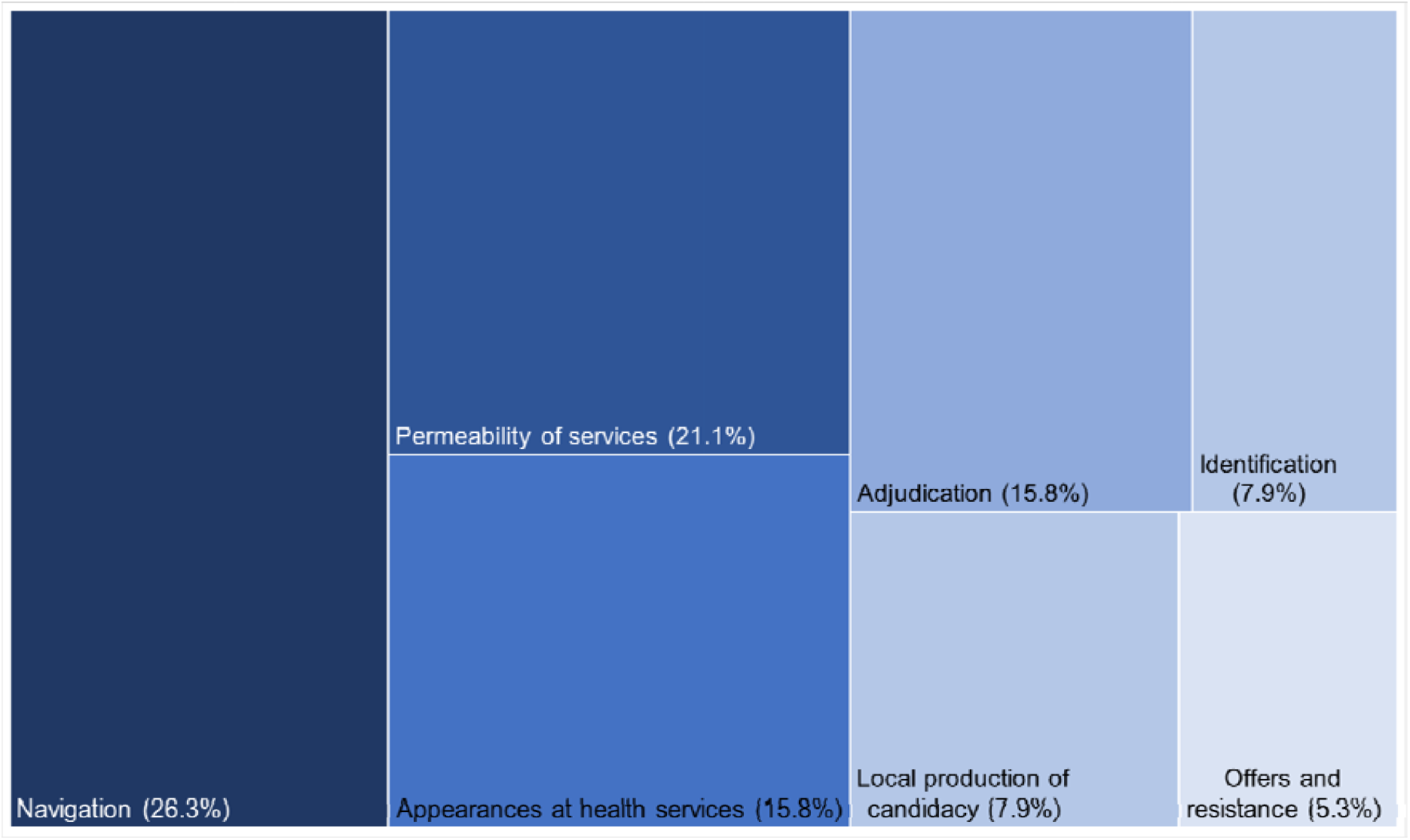
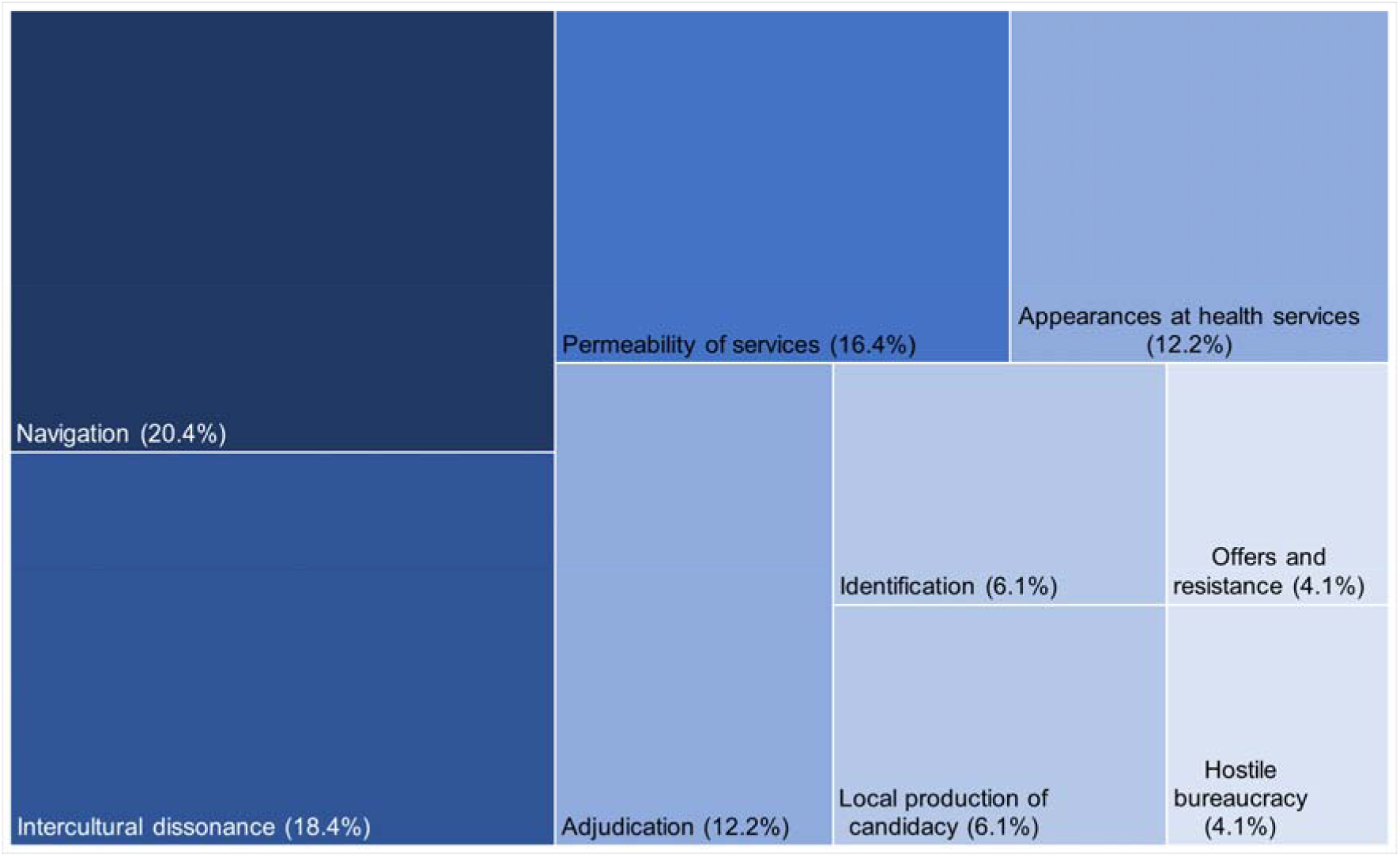
Thematic contribution to the original and extended 7+2 Candidacy Framework respectively.

## Discussion

### Main findings

This systematic review identified 51 qualitative studies documenting, across 14 HICs, maternity care-seeking experiences of more than 1,300 women from minoritised and underserved groups. Twelve sub-themes emerged across five themes: (1) Loss of dignity, autonomy, and personhood; (2) Lack of informed choice and decision-making; (3) Trust *in* and relationships *with* HCPs; (4) Differences between healthcare systems and cultures; and (5) Systemic barriers. Experiences were largely negative. While sub-themes aligned with all seven components of the Candidacy Framework, most mapped to ‘navigation’, ‘permeability of services’, and ‘appearances at health services’, highlighting shared responsibility for improving care. Two new constructs—*intercultural dissonance* and *hostile bureaucracy*— emerged, particularly affecting migrants through altered intergenerational roles and exclusionary immigration policies. The meta-ethnography provided an analytic synthesis, rendering the theory: *‘Respect, informed choice, and trust enhances candidacy whilst differences in healthcare systems, culture, and systemic barriers have the propensity to diminish it’*.

### Comparison with the literature

To our knowledge, this is the first systematic review focused exclusively on care-seeking experiences of diverse minoritised and underserved groups in HICs. Unlike our qualitative approach, most care-seeking research is quantitative, measuring attendance, visit frequency, or utilisation—often inconsistently defined ^86^ —and linking these to pregnancy outcomes. Frameworks like the social determinants of health (SDoH) model have been used to assess drivers of care-seeking, especially non-attendance,^19,25^ including socio-cultural, political, and economic factors.^19^

We build on a small number of reviews examining antenatal care among underserved groups (e.g., ethnic minorities, immigrants), which highlight complex barriers such as limited language skills, poor awareness of services, immigration and financial constraints, prior negative care experiences, and structural or organisational challenges.^6,87,88^

This review found that care-seeking experiences were largely negative. A key issue was the lack of respectful treatment—dismissed concerns, unanswered questions, and unkind interactions—which left women feeling dehumanised. ^75,78,89^ Prior research in LMICs shows that disrespectful care erodes trust and delays healthcare use. ^90,91^ For women with physical disabilities, inadequate attention to accessibility worsened this, leading to a loss of dignity. ^76^ Stigma, discrimination, and insufficient information further undermined autonomy.^55,78^ Quantitative studies also associate physical disability and one or more social risk factors to increased experiences of identity-related disrespect and reduced autonomy in maternity care. ^92,93^

Our work demonstrates barriers to healthcare-seeking in pregnancy are jointly-driven, based on how frequently our sub-themes map to factors within the Candidacy Framework^19^. Previous studies support this, identifying both system-level factors (e.g., organisational processes and system policies)^94–97^ and individual-level factors (e.g., poor doctor-patient relationship,^98^ stigmatisation,^99^ or being dismissed).^100^ Improving care engagement for underserved women requires joint negotiation and co-production of services—such as the UK’s Maternity and Neonatal Voices Partnerships (MNVPs). The limited literature speaking to the constructs of the Candidacy Framework: ‘offers and resistance’ and ‘identification’ – joint- and individual-level factors – often places blame on women for low engagement attributing it to poor health literacy,^57^ problematising their language skils,^101,102^ and further stigmatising this already marginalised population.

A unique contribution of our study is the identification of *intercultural dissonance* and *hostile bureaucracy* as additions to the Candidacy Framework, reflecting the lasting and intergenerational effects of migration on care-seeking, including during pregnancy. Events of the last decade have emphasised the underserved nature of this population which has grown exponentially in the recent past due to various humanitarian crises. Differences in healthcare systems and cultures between ‘home’ countries and ‘host’ countries, significantly shape decisions about when and how to seek care, navigate services, and act on medical advice.^69^ This can lead to an *authoritative knowledge struggle*, where contradictory information,^59,103^ may cause women to disengage from care altogether. ^44^ Additionally, psychological research highlights how generational trauma and inherited knowledge influence wellbeing and behaviour.^104^ A UK review of eight studies on asylum-seeking women identified barriers such as poor awareness of services, communication struggles, and stigma but did not explore how differing healthcare norms affect maternity experiences.^97^ Our meta-ethnographic approach, being generative and interpretive, was likely more attuned to these dynamics.

While our synthesis identifies common themes in migrant women’s experiences, the 13 HICs represented (e.g., UK, USA, Saudi Arabia) vary widely in healthcare models, migrant entitlements, and cultural expectations. For instance, healthcare fees in the US or restrictions on undocumented migrants in Europe may intensify systemic barriers compared to countries with universal access. These structural and cultural differences affect the transferability of findings, particularly in relation to how barriers manifest and are addressed. Tools such as the WHO Health Financing Progress Matrix or Migrant Integration Policy Index which examines how well a country’s health financing policies align with achieving universal coverage,^105^ and policies to integrate migrants and other marginalised groups into society^106^ respectively show marked difference between our 14 included countries. Only two countries (Sweden and Canada) ranked highly in both assessments. Such factors are likely to influence care-seeking behaviour, shaping the relevance of our findings.

Additionally, while our sample includes a diverse group of marginalised communities with several complex social risk factors, only two^54,80^ of the 51 studies consider the idea of intersectionality. It has now been well-established that individuals with multiple marginalised identities face compounded barriers to care access and utilisation,^107^ and future research is crucial to understanding these unique intersections of disadvantage.

### Strengths, limitations, and future directions

A key strength of our review is the use of meta-ethnography, allowing us to build on themes from individual studies—all of which were of moderate-to-high quality—and identify gaps in existing theory. Notably, we highlight the multi-generational impact of immigration on care-seeking as a missing component of the Candidacy Framework, supporting its expansion. Our focus on women’s care-seeking excluded perspectives of fathers, partners, non-gestational parents, providers, and policymakers. While we observed similarities across groups and countries, we may have missed group-specific or system-level differences, which we plan to explore further. A planned sub-group analysis on the pandemic’s impact was not possible due to limited studies; questions remain on how service reconfigurations and misinformation shaped care-seeking during this time and will be explored by is in future qualitative work. Future research should also examine the roles of families, professionals, and health systems, and empirically validate the proposed construct of intercultural dissonance.

## Conclusion

In HICs, maternity care-seeking is a joint responsibility between service-users and service-providers. As such, interventions to remove barriers to care-seeking should be co-produced through collaborative means between stakeholders. Efforts to improve utilisation *of,* and engagement *with,* antenatal care services should prioritise alleviating system-level barriers. We suggest an expansion of the Candidacy Framework to include two further dimensions which reflect the multigenerational effect of migration on care experience and the often hostile and precarious bureaucratic environment in which women find themselves when attempting to seek maternity care.

## Data Availability

Not applicable- this is a systematic review.

## Abbreviations

UK: United Kingdom
HIC: High-income country
LMIC: Low- and middle-income country
HCP: Healthcare professional
SES: Socioeconomic status
USA: United States of America
WHO: World Health Organization
SDoH: Social determinants of Health
CASP: Critical Appraisal Skills Programme
IPV: Intimate partner violence

## Declarations

### Patient and Public Engagement and Involvement (PPIE)

The study was reviewed by an established PPIE group at the NIHR Applied Research Collaboration [ARC] in South London, to ensure coherence with service users’ lived experience and ensuring relevance of the research question and search criteria. The study PPIE advisory group (YB,MP) have been involved in editing and review of this manuscript.

### Ethics approval and consent to participate

Not applicable.

### Consent to publication

Not applicable.

### Availability of data and materials

Not applicable.

### Competing Interests

None to declare.

### Funding

Tisha Dasgupta is in receipt of an Economic and Social Research Council [ESRC] doctoral training fellowship from the London Interdisciplinary Social Science Doctoral Training Partnership [LISS DTP], (ES/P000703/1). Hannah Rayment-Jones is funded by a NIHR Advanced Fellowship (NIHR 303183).

### Author contributions

Conceptualization: TD, HRJ, LAM, SAS; Data curation: TD, HRJ; Formal analysis: TD, GH, HRJ; Funding acquisition: TD; Investigation: TD, HRJ, SAS, LAM; Methodology: TD, SAS; Project administration: TD; Software: TD, GH; Resources: LAM, HRJ, SAS; Supervision: LAM, HRJ, SAS; Validation: TD, HRJ, SAS, LAM, YB, MP; Visualization: TD, HRJ; Writing – original draft: TD; Writing – review & editing: SAS, LAM, HRJ, GH, YB, MP

### Authors’ information

The ENGAGE Study, to which this systematic review contributes, has been adopted by the National Institute for Health and Care Research Applied Research Collaboration South London [NIHR ARC South London] at King’s College Hospital NHS Foundation Trust. The views expressed are those of the authors and not necessarily those of the NIHR or the Department of Health and Social Care.

During the preparation of this work the author(s) used ChatGPT in order to make the manuscript more concise and reduce words. After using this tool/service, the author(s) reviewed and edited the content as needed and take(s) full responsibility for the content of the publication.

## Notes

### Competing Interest Statement

The authors have declared no competing interest.

## References

1. World Health Organization. WHO recommendations on antenatal care for a positive pregnancy experience. 2016.

2. Tunçalp, Pena-Rosas JP, Lawrie T, Bucagu M, Oladapo OT, Portela A, et al. WHO recommendations on antenatal care for a positive pregnancy experience—going beyond survival. BJOG. 2017 May 1;124(6):860–2.

3. Stacey T, Thompson JMD, Mitchell EA, Zuccollo JM, Ekeroma AJ, McCowan LME. Antenatal care, identification of suboptimal fetal growth and risk of late stillbirth: Findings from the Auckland Stillbirth Study. Australian and New Zealand Journal of Obstetrics and Gynaecology [Internet]. 2012 Jun 1 [cited 2024 May 22];52(3):242–7. Available from: https://onlinelibrary.wiley.com/doi/full/10.1111/j.1479-828X.2011.01406.x

4. Kuhnt J, Vollmer S. Antenatal care services and its implications for vital and health outcomes of children: Evidence from 193 surveys in 69 low-income and middle-income countries. BMJ Open. 2017 Nov 1;7(11).

5. Sarikhani Y, Najibi SM, Razavi Z. Key barriers to the provision and utilization of maternal health services in low-and lower-middle-income countries; a scoping review. BMC Womens Health. 2024 Dec 1;24(1).

6. Downe S, Finlayson K, Tunçalp Ö, Gülmezoglu AM. Provision and uptake of routine antenatal services: A qualitative evidence synthesis. Cochrane Database of Systematic Reviews. 2019 Jun 12;2019(6).

7. Mamrath S, Greenfield M, Turienzo CF, Fallon V, Silverio SA. Experiences of postpartum anxiety during the COVID-19 pandemic: A mixed methods study and demographic analysis. PLoS One [Internet]. 2024 Mar 1 [cited 2024 Jul 17];19(3):e0297454. Available from: https://journals.plos.org/plosone/article?id=10.1371/journal.pone.0297454

8. Redshaw M, Heikkila K. Delivered with care: Delivered with care: a national survey of women’s experience of maternity care 2010. 2010.

9. Escañuela Sánchez T, Linehan L, O’Donoghue K, Byrne M, Meaney S. Facilitators and barriers to seeking and engaging with antenatal care in high-income countries: A meta-synthesis of qualitative research. Health Soc Care Community. 2022 Nov 1;30(6):e3810–28.

10. Peterson L, Bridle L, Dasgupta T, Easter A, Ghobrial S, Ishlek I, et al. Oscillating autonomy: A grounded theory study of women’s experiences of COVID-19 infection during pregnancy, labour and birth, and the early postnatal period. BMC Pregnancy Childbirth [Internet]. 2024 Jul 8 [cited 2024 Jul 17]; Available from: https://kclpure.kcl.ac.uk/portal/en/publications/oscillating-autonomy-a-grounded-theory-study-of-womens-experience

11. Jackson L, Greenfield M, Payne E, Burgess K, Oza M, Storey C, et al. A consensus statement on perinatal mental health during the COVID-19 pandemic and recommendations for post-pandemic recovery and re-build. Front Glob Womens Health. 2024 Feb 21;5:1347388.

12. Silverio SA, Harris EJ, Jackson L, Fallon V, Easter A, Dadelszen P von, et al. Freedom for some, but not for Mum: the reproductive injustice associated with pandemic ‘Freedom Day’ for perinatal women in the United Kingdom. Front Public Health. 2024;12.

13. Montgomery E, De Backer K, Easter A, Magee LA, Sandall J, Silverio SA. Navigating uncertainty alone: A grounded theory analysis of women’s psycho-social experiences of pregnancy and childbirth during the COVID-19 pandemic in London. Women and Birth. 2023 Feb 1;36(1):e106–17.

14. Jardine J, Relph S, Magee LA, von Dadelszen P, Morris E, Ross-Davie M, et al. Maternity services in the UK during the coronavirus disease 2019 pandemic: a national survey of modifications to standard care. BJOG [Internet]. 2021 Apr 1 [cited 2023 Jul 7];128(5):880–9. Available from: https://onlinelibrary.wiley.com/doi/full/10.1111/1471-0528.16547

15. Bridle L, Walton L, van der Vord T, Adebayo O, Hall S, Finlayson E, et al. Supporting Perinatal Mental Health and Wellbeing during COVID-19. Int J Environ Res Public Health. 2022 Feb 4;19(3):1777.

16. Dasgupta T, Horgan G, Peterson L, Mistry HD, Balls E, Wilson M, et al. Women’s experiences of maternity care in the United Kingdom during the COVID-19 pandemic: A follow-up systematic review and qualitative evidence synthesis. Women and Birth. 2024;37(3).

17. Jackson L, De Pascalis L, Harrold JA, Fallon V, Silverio SA. Postpartum women’s experiences of social and healthcare professional support during the COVID-19 pandemic: A recurrent cross-sectional thematic analysis. Women and Birth. 2022 Sep 1;35(5):511–20.

18. Silverio SA, De Backer K, Easter A, von Dadelszen P, Magee LA, Sandall J. Women’s experiences of maternity service reconfiguration during the COVID-19 pandemic: A qualitative investigation. Midwifery. 2021 Nov 1;102:103116.

19. Dixon-Woods M, Cavers D, Agarwal S, Annandale E, Arthur A, Harvey J, et al. Conducting a critical interpretive synthesis of the literature on access to healthcare by vulnerable groups. BMC Med Res Methodol [Internet]. 2006 Jul 26 [cited 2022 Nov 4];6(1):1–13. Available from: https://bmcmedresmethodol.biomedcentral.com/articles/10.1186/1471-2288-6-35

20. Tookey S, Renzi C, Waller J, Von Wagner C, Whitaker KL. Using the candidacy framework to understand how doctor-patient interactions influence perceived eligibility to seek help for cancer alarm symptoms: A qualitative interview study. BMC Health Serv Res [Internet]. 2018 Dec 4 [cited 2025 Jun 13];18(1):1–8. Available from: https://bmchealthservres.biomedcentral.com/articles/10.1186/s12913-018-3730-5

21. Koehn S. Negotiating candidacy: ethnic minority seniors’ access to care. Ageing Soc [Internet]. 2009 [cited 2025 Jun 13];29(4):585–608. Available from: https://www.cambridge.org/core/journals/ageing-and-society/article/abs/negotiating-candidacy-ethnic-minority-seniors-access-to-care/EF7CC467B08637DDE2D72680FE5EA04B

22. Mackenzie M, Conway E, Hastings A, Munro M, O’Donnell C. Is ‘Candidacy’ a Useful Concept for Understanding Journeys through Public Services? A Critical Interpretive Literature Synthesis. Soc Policy Adm [Internet]. 2013 Dec 1 [cited 2025 Jun 13];47(7):806–25. Available from: /doi/pdf/10.1111/j.1467-9515.2012.00864.x

23. Liberati E, Richards N, Parker J, Willars J, Scott D, Boydell N, et al. Qualitative study of candidacy and access to secondary mental health services during the COVID-19 pandemic. Soc Sci Med [Internet]. 2022 Mar 1 [cited 2025 Jun 13];296:114711. Available from: https://www.sciencedirect.com/science/article/pii/S0277953622000144#bib8

24. Rayment-Jones H, Silverio SA, Harris J, Harden A, Sandall J. Project 20: Midwives’ insight into continuity of care models for women with social risk factors: what works, for whom, in what circumstances, and how. Midwifery. 2020 May 1;84:102654.

25. Hinton L, Kuberska K, Dakin F, Boydell N, Martin G, Draycott T, et al. A qualitative study of the dynamics of access to remote antenatal care through the lens of candidacy. J Health Serv Res Policy. 2023 Oct 1;28(4):222–32.

26. Page MJ, McKenzie JE, Bossuyt PM, Boutron I, Hoffmann TC, Mulrow CD, et al. The PRISMA 2020 statement: an updated guideline for reporting systematic reviews. Syst Rev [Internet]. 2021 Dec 1 [cited 2023 Sep 16];10(1). Available from: https://pubmed.ncbi.nlm.nih.gov/33781348/

27. Tisha Dasgupta - LISS DTP [Internet]. [cited 2025 Jan 17]. Available from: https://liss-dtp.ac.uk/students/tisha-dasgupta/

28. Ouzzani M, Hammady H, Fedorowicz Z, Elmagarmid A. Rayyan—a web and mobile app for systematic reviews. Syst Rev. 2016 Dec 5;5(1):210.

29. Critical Appraisal Skills Programme. CASP Qualitative Checklist [Internet]. 2018 [cited 2024 May 22]. Available from: https://casp-uk.net/checklists/casp-qualitative-studies-checklist-fillable.pdf

30. W.Noblit G, DwightHare R. Meta-Ethnography. Meta-Ethnography. 1988 May 18;

31. Sattar R, Lawton R, Panagioti M, Johnson J. Meta-ethnography in healthcare research: a guide to using a meta-ethnographic approach for literature synthesis. [cited 2024 May 13]; Available from: 10.1186/s12913-020-06049-w

32. Britten N, Campbell R, Pope C, Donovan J, Morgan M, Pill R. Using meta ethnography to synthesise qualitative research: A worked example. J Health Serv Res Policy [Internet]. 2002 Oct [cited 2025 May 10];7(4):209–15. Available from: /doi/pdf/10.1258/135581902320432732?download=true

33. Malpass A, Shaw A, Sharp D, Walter F, Feder G, Ridd M, et al. “Medication career” or “Moral career”? The two sides of managing antidepressants: A meta-ethnography of patients’ experience of antidepressants. Soc Sci Med [Internet]. 2009 Jan 1 [cited 2025 May 10];68(1):154–68. Available from: https://www.sciencedirect.com/science/article/pii/S0277953608005133?via%3Dihub#sec2

34. Barkensjö M, Greenbrook JTV, Rosenlundh J, Ascher H, Elden H. The need for trust and safety inducing encounters: A qualitative exploration of women’s experiences of seeking perinatal care when living as undocumented migrants in Sweden. BMC Pregnancy Childbirth [Internet]. 2018 Jun 7 [cited 2024 Jul 17];18(1):1–17. Available from: https://bmcpregnancychildbirth.biomedcentral.com/articles/10.1186/s12884-018-1851-9

35. Tesfalul MA, Feuer SK, Castillo E, Coleman-Phox K, O’Leary A, Kuppermann M. Patient and provider perspectives on preterm birth risk assessment and communication. Patient Educ Couns [Internet]. 2021 Nov 1 [cited 2024 Jul 17];104(11):2814–23. Available from: https://pubmed.ncbi.nlm.nih.gov/33892976/

36. Ayers BL, Purvis RS, Bing WI, Rubon-Chutaro J, Hawley NL, Delafield R, et al. Structural and Socio-cultural Barriers to Prenatal Care in a US Marshallese Community. Matern Child Health J [Internet]. 2018 Jul 1 [cited 2024 Jul 17];22(7):1067–76. Available from: https://link.springer.com/article/10.1007/s10995-018-2490-5

37. McLemore MR, Altman MR, Cooper N, Williams S, Rand L, Franck L. Health care experiences of pregnant, birthing and postnatal women of color at risk for preterm birth. Soc Sci Med. 2018 Mar 1;201:127–35.

38. Vang ZM, Gagnon R, Lee T, Jimenez V, Navickas A, Pelletier J, et al. Interactions Between Indigenous Women Awaiting Childbirth Away From Home and Their Southern, Non-Indigenous Health Care Providers. 10.1177/1049732318792500 [Internet]. 2018 Aug 10 [cited 2024 Jul 17];28(12):1858–70. Available from: https://journals.sagepub.com/doi/10.1177/1049732318792500

39. Holten L, Hollander M, de Miranda E. When the Hospital Is No Longer an Option: A Multiple Case Study of Defining Moments for Women Choosing Home Birth in High-Risk Pregnancies in The Netherlands. Qual Health Res [Internet]. 2018 Oct 1 [cited 2024 Jul 17];28(12):1883–96. Available from: https://journals.sagepub.com/doi/10.1177/1049732318791535

40. Origlia Ikhilor P, Hasenberg G, Kurth E, Asefaw F, Pehlke-Milde J, Cignacco E. Communication barriers in maternity care of allophone migrants: Experiences of women, healthcare professionals, and intercultural interpreters. J Adv Nurs [Internet]. 2019 Oct 1 [cited 2024 Jul 17];75(10):2200–10. Available from: https://onlinelibrary.wiley.com/doi/full/10.1111/jan.14093

41. Sami J, Lötscher KCQ, Eperon I, Gonik L, De Tejada BM, Epiney M, et al. Giving birth in Switzerland: A qualitative study exploring migrant women’s experiences during pregnancy and childbirth in Geneva and Zurich using focus groups. Reprod Health [Internet]. 2019 Jul 22 [cited 2024 Jul 17];16(1):1–9. Available from: https://reproductive-health-journal.biomedcentral.com/articles/10.1186/s12978-019-0771-0

42. Spangaro J, Koziol-McLain J, Rutherford A, Zwi AB. “Made Me Feel Connected”: A Qualitative Comparative Analysis of Intimate Partner Violence Routine Screening Pathways to Impact. 10.1177/1077801219830250 [Internet]. 2019 Mar 14 [cited 2024 Jul 17];26(3–4):334–58. Available from: https://journals.sagepub.com/doi/10.1177/1077801219830250

43. Holden G, Corter AL, Hatters-Friedman S, Soosay I. Brief Report. A qualitative study of maternal mental health services in New Zealand: Perspectives of Māori and Pacific mothers and midwives. Asia-Pacific Psychiatry [Internet]. 2019 Jun 1 [cited 2024 Jul 17];12(2):e12369. Available from: https://onlinelibrary.wiley.com/doi/full/10.1111/appy.12369

44. Johnsen H, Christensen U, Juhl M, Villadsen SF. Contextual Factors Influencing the MAMAACT Intervention: A Qualitative Study of Non-Western Immigrant Women’s Response to Potential Pregnancy Complications in Everyday Life. International Journal of Environmental Research and Public Health 2020, Vol 17, Page 1040 [Internet]. 2020 Feb 6 [cited 2024 Jul 17];17(3):1040. Available from: https://www.mdpi.com/1660-4601/17/3/1040/htm

45. Decker MJ, Pineda N, Gutmann-Gonzalez A, Brindis CD. Youth-centered maternity care: a binational qualitative comparison of the experiences and perspectives of Latina adolescents and healthcare providers. BMC Pregnancy Childbirth [Internet]. 2021 Dec 1 [cited 2024 Jul 17];21(1):1–12. Available from: https://bmcpregnancychildbirth.biomedcentral.com/articles/10.1186/s12884-021-03831-4

46. Bains S, Skråning S, Sundby J, Vangen S, Sørbye IK, Lindskog B V. Challenges and barriers to optimal maternity care for recently migrated women - a mixed-method study in Norway. BMC Pregnancy Childbirth [Internet]. 2021 Dec 1 [cited 2024 Jul 17];21(1):1–14. Available from: https://bmcpregnancychildbirth.biomedcentral.com/articles/10.1186/s12884-021-04131-7

47. Hjelm K, Bard K, Apelqvist J. A qualitative study of developing beliefs about health, illness and healthcare in migrant African women with gestational diabetes living in Sweden. BMC Womens Health [Internet]. 2018 Feb 5 [cited 2024 Jul 17];18(1):1–14. Available from: https://bmcwomenshealth.biomedcentral.com/articles/10.1186/s12905-018-0518-z

48. Nottingham-Jones J, Simmonds JG, Snell TL. First-time mothers’ experiences of preparing for childbirth at advanced maternal age. Midwifery. 2020 Jul 1;86:102558.

49. Utne R, Antrobus-Johannessen CL, Aasheim V, Aaseskjær K, Vik ES. Somali women’s experiences of antenatal care: A qualitative interview study. Midwifery. 2020 Apr 1;83:102656.

50. Bitar D, Oscarsson M. Arabic-speaking women’s experiences of communication at antenatal care in Sweden using a tablet application—Part of development and feasibility study. Midwifery. 2020 May 1;84:102660.

51. Oza-Frank R, Conrey E, Bouchard J, Shellhaas C, Weber MB. Healthcare Experiences of Low-Income Women with Prior Gestational Diabetes. Matern Child Health J [Internet]. 2018 Jul 1 [cited 2024 Jul 17];22(7):1059–66. Available from: https://link.springer.com/article/10.1007/s10995-018-2489-y

52. Byatt N, Straus J, Stopa A, Biebel K, Mittal L, Simas TAM. Massachusetts Child Psychiatry Access Program for Moms: Utilization and Quality Assessment. Obstetrics and Gynecology [Internet]. 2018 [cited 2024 Jul 17];132(2):345–53. Available from: https://journals.lww.com/greenjournal/fulltext/2018/08000/massachusetts_child_psychiatry_access_program_for.11.aspx

53. Drago MJ, Guillén U, Schiaratura M, Batza J, Zygmunt A, Mowes A, et al. Constructing a Culturally Informed Spanish Decision-Aid to Counsel Latino Parents Facing Imminent Extreme Premature Delivery. Matern Child Health J [Internet]. 2018 Jul 1 [cited 2024 Jul 17];22(7):950–7. Available from: https://link.springer.com/article/10.1007/s10995-018-2471-8

54. Daoud N, Abu-Hamad S, Berger-Polsky A, Davidovitch N, Orshalimy S. Mechanisms for racial separation and inequitable maternal care in hospital maternity wards. Soc Sci Med. 2022 Jan 1;292:114551.

55. Wah YYE, McGill M, Wong J, Ross GP, Harding AJ, Krass I. Self-management of gestational diabetes among Chinese migrants: A qualitative study. Women and Birth. 2019 Feb 1;32(1):e17–23.

56. Farewell C V., Jewell J, Walls J, Leiferman JA. A Mixed-Methods Pilot Study of Perinatal Risk and Resilience During COVID-19. J Prim Care Community Health [Internet]. 2020 Jul 16 [cited 2024 Jul 17];11. Available from: https://journals.sagepub.com/doi/10.1177/2150132720944074

57. Savory NA, Hannigan B, Sanders J. Women’s experience of mild to moderate mental health problems during pregnancy, and barriers to receiving support. Midwifery. 2022 May 1;108:103276.

58. Alanazy W, Rance J, Brown A. Exploring maternal and health professional beliefs about the factors that affect whether women in Saudi Arabia attend antenatal care clinic appointments. Midwifery. 2019 Sep 1;76:36–44.

59. Henry J, Beruf C, Fischer T. Access to Health Care for Pregnant Arabic-Speaking Refugee Women and Mothers in Germany. Qualitatiive Health Research [Internet]. 2019 Sep 18 [cited 2024 Jul 17];30(3):437–47. Available from: https://journals.sagepub.com/doi/10.1177/1049732319873620

60. Barkin JL, Bloch JR, Smith KER, Telliard SN, McGreal A, Sikes C, et al. Knowledge of and Attitudes Toward Perinatal Home Visiting in Women with High-Risk Pregnancies. J Midwifery Womens Health [Internet]. 2021 Mar 1 [cited 2024 Jul 17];66(2):227–32. Available from: https://onlinelibrary.wiley.com/doi/full/10.1111/jmwh.13204

61. Frederiksen MS, Schmied V, Overgaard C. Living With Fear: Experiences of Danish Parents in Vulnerable Positions During Pregnancy and in the Postnatal Period. Qual Health Res [Internet]. 2021 Jan 10 [cited 2024 Jul 17];31(3):564–77. Available from: https://journals.sagepub.com/doi/10.1177/1049732320978206

62. Frederiksen MS, Schmied V, Overgaard C. Supportive encounters during pregnancy and the postnatal period: An ethnographic study of care experiences of parents in a vulnerable position. J Clin Nurs [Internet]. 2021 Aug 1 [cited 2024 Jul 17];30(15–16):2386–98. Available from: https://onlinelibrary.wiley.com/doi/full/10.1111/jocn.15778

63. Reid CN, Fryer K, Cabral N, Marshall J. Health care system barriers and facilitators to early prenatal care among diverse women in Florida. Birth [Internet]. 2021 Sep 1 [cited 2024 Jul 17];48(3):416–27. Available from: https://onlinelibrary.wiley.com/doi/full/10.1111/birt.12551

64. (Fontein)Kuipers YJ, Mestdagh E. The experiential knowledge of migrant women about vulnerability during pregnancy: A woman-centred mixed-methods study. Women and Birth. 2022 Feb 1;35(1):70–9.

65. Mule V, Reilly NM, Schmied V, Kingston D, Austin MP V. Why do some pregnant women not fully disclose at comprehensive psychosocial assessment with their midwife? Women and Birth. 2022 Feb 1;35(1):80–6.

66. McClellan C, Madler B. Lived Experiences of Mongolian Immigrant Women Seeking Perinatal Care in the United States. 10.1177/10436596221091689 [Internet]. 2022 May 10 [cited 2024 Jul 17];33(5):594–602. Available from: https://journals.sagepub.com/doi/10.1177/10436596221091689?url_ver=Z39.88-2003&rfr_id=ori%3Arid%3Acrossref.org&rfr_dat=cr_pub++0pubmed

67. Njenga A. Somali Refugee Women’s Experiences and Perceptions of Western Health Care. Journal of Transcultural Nursing [Internet]. 2022 Oct 5 [cited 2024 Jul 17];34(1):8–13. Available from: https://journals.sagepub.com/doi/10.1177/10436596221125893

68. Mehrara L, Olaug Gjernes TK, Young S. Immigrant women’s experiences with Norwegian maternal health services: implications for policy and practice. Int J Qual Stud Health Well-being [Internet]. 2022 Dec 31 [cited 2024 Jul 17];17(1). Available from: https://www.tandfonline.com/doi/abs/10.1080/17482631.2022.2066256

69. Goodwin L, Hunter B, Jones A. The midwife–woman relationship in a South Wales community: Experiences of midwives and migrant Pakistani women in early pregnancy. Health Expect [Internet]. 2018 Feb 1 [cited 2024 Jul 17];21(1):347. Available from: /pmc/articles/PMC5750740/

70. Wilson R, Paterson P, Larson HJ. Strategies to improve maternal vaccination acceptance. BMC Public Health [Internet]. 2019 Mar 25 [cited 2024 Jul 17];19(1). Available from: /pmc/articles/PMC6434850/

71. Naylor Smith J, Taylor B, Shaw K, Hewison A, Kenyon S. “I didn’t think you were allowed that, they didn’t mention that.” A qualitative study exploring women’s perceptions of home birth. BMC Pregnancy Childbirth [Internet]. 2018 Apr 18 [cited 2024 Jul 17];18(1):1–11. Available from: https://bmcpregnancychildbirth.biomedcentral.com/articles/10.1186/s12884-018-1733-1

72. Husain F, Powys VR, White E, Jones R, Goldsmith LP, Heath PT, et al. COVID-19 vaccination uptake in 441 socially and ethnically diverse pregnant women. PLoS One [Internet]. 2022 Aug 1 [cited 2024 Jul 17];17(8). Available from: /pmc/articles/PMC9385003/

73. Stacey T, Haith-Cooper M, Almas N, Kenyon C. An exploration of migrant women’s perceptions of public health messages to reduce stillbirth in the UK: a qualitative study. BMC Pregnancy Childbirth [Internet]. 2021 Dec 1 [cited 2024 Jul 17];21(1):1–9. Available from: https://bmcpregnancychildbirth.biomedcentral.com/articles/10.1186/s12884-021-03879-2

74. Gong Q (Sarah) S, Bharj K. A qualitative study of the utilisation of digital resources in pregnant Chinese migrant women’s maternity care in northern England. Midwifery. 2022 Dec 1;115:103493.

75. Balaam MC, Thomson G. Building capacity and wellbeing in vulnerable/marginalised mothers: A qualitative study. Women and Birth. 2018 Oct 1;31(5):e341–7.

76. Hall J, Hundley V, Collins B, Ireland J. Dignity and respect during pregnancy and childbirth: A survey of the experience of disabled women. BMC Pregnancy Childbirth [Internet]. 2018 Aug 13 [cited 2024 Jul 17];18(1):1–13. Available from: https://bmcpregnancychildbirth.biomedcentral.com/articles/10.1186/s12884-018-1950-7

77. Gordon ACT, Burr J, Lehane D, Mitchell C. Influence of past trauma and health interactions on homeless women’s views of perinatal care: a qualitative study. The British Journal of General Practice [Internet]. 2019 [cited 2024 Jul 17];69(688):e760. Available from: /pmc/articles/PMC6733590/

78. McLeish J, Redshaw M. Maternity experiences of mothers with multiple disadvantages in England: A qualitative study. Women and Birth. 2019 Apr 1;32(2):178–84.

79. George EK, Dominique S, Irie W, Edmonds JK. “It’s my Home away from Home:” A hermeneutic phenomenological study exploring decision-making experiences of choosing a freestanding birth centre for perinatal care. Midwifery [Internet]. 2024 Dec 1 [cited 2025 May 24];139:104164. Available from: https://www.sciencedirect.com/science/article/pii/S026661382400247X?via%3Dihub

80. Parker G, Miller S, Ker A, Baddock S, Kerekere E, Veale J. “Let All Identities Bloom, Just Let Them Bloom”: Advancing Trans-Inclusive Perinatal Care Through Intersectional Analysis. Qual Health Res [Internet]. 2025 Apr 1 [cited 2025 May 24];35(4–5):403–17. Available from: https://scholar.google.com/scholar_url?url=https://journals.sagepub.com/doi/pdf/10.1177/10497323241309590&hl=en&sa=T&oi=ucasa&ct=usl&ei=JQ8yaIXxHeW16rQPsqVy&scisig=AAZF9b_Twrdk22B-j7aJ0SceLajy

81. Faulks F, Shafiei T, Mogren I, Edvardsson K. “It’s just too far…”: A qualitative exploration of the barriers and enablers to accessing perinatal care for rural Australian women. Women and Birth [Internet]. 2024 Nov 1 [cited 2025 May 24];37(6):101809. Available from: https://www.sciencedirect.com/science/article/pii/S1871519224002695?via%3Dihub

82. Nechaeva E, Kharkova O, Postoev V, Grjibovski AM, Darj E, Odland JØ. Awareness of postpartum depression among midwives and pregnant women in Arkhangelsk, Arctic Russia. Glob Health Action [Internet]. 2024 [cited 2025 May 24];17(1):2354008. Available from: https://pmc.ncbi.nlm.nih.gov/articles/PMC11149570/

83. Pierce P, Whitten M, Hillman S. The impact of digital healthcare on vulnerable pregnant women: A review of the use of the MyCare app in the maternity department at a central London tertiary unit. Front Digit Health. 2023 Apr 21;5:1155708.

84. Yuill C, Sinesi A, Meades R, Williams LR, Delicate A, Cheyne H, et al. Women’s experiences and views of routine assessment for anxiety in pregnancy and after birth: A qualitative study. Br J Health Psychol [Internet]. 2024 Nov 1 [cited 2025 May 24];29(4). Available from: https://pubmed.ncbi.nlm.nih.gov/38955505/

85. Tuckett A. Rules, Paper, Status Migrants and Precarious Bureaucracy in Contemporary Italy. Stanford University Press; 2018.

86. Mackian S, Bedri N, Lovel H. Up the garden path and over the edge: where might health-seeking behaviour take us? Health Policy Plan [Internet]. 2004 May 1 [cited 2022 Jan 23];19(3):137–46. Available from: https://academic.oup.com/heapol/article/19/3/137/774112

87. Sharma E, Tseng PC, Harden A, Li L, Puthussery S. Ethnic minority women’s experiences of accessing antenatal care in high income European countries: a systematic review. BMC Health Serv Res. 2023;23:612.

88. Higginbottom G, Evans C, Morgan M, Bharj K, Eldridge J, Hussain B. Experience of and access to maternity care in the United Kingdom (UK) by immigrant women: a narrative synthesis systematic review. 2019;

89. Silverio SA, Varman N, Barry Z, Khazaezadeh N, Rajasingam D, Magee LA, et al. Inside the ‘imperfect mosaic’: Minority ethnic women’s qualitative experiences of race and ethnicity during pregnancy, childbirth, and maternity care in the United Kingdom. BMC Public Health. 2023 Dec 21;23(1):2555.

90. Puthussery S, Bayih WA, Brown H, Aborigo RA. Promoting a global culture of respectful maternity care. BMC Pregnancy Childbirth [Internet]. 2023 Dec 1 [cited 2024 Sep 9];23(1):1–3. Available from: https://bmcpregnancychildbirth.biomedcentral.com/articles/10.1186/s12884-023-06118-y

91. Bohren MA, Hunter EC, Munthe-Kaas HM, Souza JP, Vogel JP, Gülmezoglu AM. Facilitators and barriers to facility-based delivery in low- and middle-income countries: A qualitative evidence synthesis. Reprod Health [Internet]. 2014 Sep 19 [cited 2024 Sep 9];11(1):1–17. Available from: https://link.springer.com/articles/10.1186/1742-4755-11-71

92. Glover A, Holman C, Boise P. Patient-centered respectful maternity care: a factor analysis contextualizing marginalized identities, trust, and informed choice. BMC Pregnancy Childbirth [Internet]. 2024 Dec 1 [cited 2024 Sep 9];24(1):1–13. Available from: https://bmcpregnancychildbirth.biomedcentral.com/articles/10.1186/s12884-024-06491-2

93. Rayment-Jones H, Dalrymple K, Harris JM, Harden A, Parslow E, Georgi T, et al. Project20: maternity care mechanisms that improve access and engagement for women with social risk factors in the UK – a mixed-methods, realist evaluation. BMJ Open. 2023 Feb 7;13(2):e064291.

94. Stanton ME, Higgs ES, Koblinsky M. Investigating Financial Incentives for Maternal Health: An Introduction. J Health Popul Nutr [Internet]. 2013 [cited 2024 Sep 9];31(4 Suppl 2):S1. Available from: /pmc/articles/PMC4021699/

95. Whitaker KL, Macleod U, Winstanley K, Scott SE, Wardle J. Help seeking for cancer ‘alarm’ symptoms: a qualitative interview study of primary care patients in the UK. British Journal of General Practice [Internet]. 2015 Feb 1 [cited 2024 Sep 9];65(631):e96–105. Available from: https://bjgp.org/content/65/631/e96

96. Richard L, Furler J, Densley K, Haggerty J, Russell G, Levesque JF, et al. Equity of access to primary healthcare for vulnerable populations: The IMPACT international online survey of innovations. Int J Equity Health [Internet]. 2016 Apr 12 [cited 2024 Sep 9];15(1):1–20. Available from: https://link.springer.com/articles/10.1186/s12939-016-0351-7

97. McKnight P, Goodwin L, Kenyon S. A systematic review of asylum-seeking women’s views and experiences of UK maternity care. Midwifery. 2019 Oct 1;77:16–23.

98. Brennan N, Barnes R, Calnan M, Corrigan O, Dieppe P, Entwistle V. Trust in the health-care provider–patient relationship: a systematic mapping review of the evidence base. International Journal for Quality in Health Care [Internet]. 2013 Dec 1 [cited 2024 Sep 9];25(6):682–8. Available from: 10.1093/intqhc/mzt063

99. Earnshaw VA, Quinn DM. The Impact of Stigma in Healthcare on People Living with Chronic Illnesses. 10.1177/1359105311414952 [Internet]. 2011 Jul 28 [cited 2024 Sep 9];17(2):157–68. Available from: https://journals.sagepub.com/doi/10.1177/1359105311414952

100. Barry CA, Stevenson FA, Britten N, Barber N, Bradley CP. Giving voice to the lifeworld. More humane, more effective medical care? A qualitative study of doctor–patient communication in general practice. Soc Sci Med. 2001 Aug 1;53(4):487–505.

101. Bridle L, Bassett S, Silverio SA. “We couldn’t talk to her”: a qualitative exploration of the experiences of UK midwives when navigating women’s care without language. Int J Hum Rights Healthc. 2021 Oct 25;14(4):359–73.

102. Rayment-Jones H, Harris J, Harden A, Silverio SA, Turienzo CF, Sandall J. Project20: interpreter services for pregnant women with social risk factors in England: what works, for whom, in what circumstances, and how? Int J Equity Health. 2021 Dec 24;20(1):233.

103. Khan Z, Vowles Z, Fernandez Turienzo C, Barry Z, Brigante L, Downe S, et al. Targeted health and social care interventions for women and infants who are disproportionately impacted by health inequalities in high-income countries: a systematic review. Int J Equity Health. 2023 Jul 11;22(1):131.

104. Wolynn M. It Didn’t Start With You: How inherited family trauma shapes who we are and how to end the cycle. reprint. Random House; 2022.

105. World Health Organization. Health Financing Progress Matrix [Internet]. [cited 2025 Jun 13]. Available from: https://www.who.int/teams/health-financing-and-economics/health-financing/diagnostics/health-financing-progress-matrix

106. Solano G, Huddleston T. Migrant Integration Policy Index. 2020.

107. Vohra-Gupta S, Petruzzi L, Jones C, Cubbin C. An Intersectional Approach to Understanding Barriers to Healthcare for Women. J Community Health [Internet]. 2022 Feb 1 [cited 2025 Jun 13];48(1):89. Available from: https://pmc.ncbi.nlm.nih.gov/articles/PMC9589537/

