## Supplementary appendix for "Understanding care-seeking of pregnant women from underserved groups: A systematic review and meta-ethnography"

### Figures

| **No.** | **Title** | **Page Number** |
| --- | --- | --- |
| S1 | PRISMA Flow chart of study selection process | 1 |

**Figure S1. PRISMA Flow chart of study selection process**

**Identification of studies via databases and registers**

Records removed *before screening*:

Duplicate records removed

(n = 605)

Update (n= 73)

Records identified from:

Databases (n = 3098)

[SCOPUS, MEDLINE, EMBASE, CINAHL, Global Health, PsychINFO, MIDRIS]

Registers (n=0 new records)

Update (n= 284)

**Identification**

Records excluded based on title and abstract

(n = 2425)

Update (n= 203)

Records screened

(n = 2493)

Update (n= 211)

**Screening**

Reports not retrieved/ wrong publication type

(n = 0)

Update (n= 0)

Full-text reports sought for retrieval (n = 68)

Update (n= 8)

Reports excluded: (n=24)

Wrong study design (n = 4)

Not relevant to review aim (n = 15)

Wrong population (n=5)

Update (n= 2)

Not relevant to review aim (n=2)

Reports assessed for eligibility

(n = 68)

Update (n= 8)

Studies included in review

(n = 45)

Update (n= 6)

**Included**

*Adapted From:*  Page MJ, McKenzie JE, Bossuyt PM, Boutron I, Hoffmann TC, Mulrow CD, et al. The PRISMA 2020 statement: an updated guideline for reporting systematic reviews. BMJ 2021;372:n71. doi: 10.1136/bmj.n71

### Tables

| **Tables** | **Title** | **Page Number** |
| --- | --- | --- |
| S1 | PRISMA checklist for reporting systematic reviews | 3 |
| S2 | Search strategy | 6 |
| S3 | Details of included studies | 9 |
| S4 | Quality appraisal of included studies | 26 |
| S5 | Mapping of sub-themes to Candidacy components | 29 |
| S6 | An extended Candidacy framework | 30 |

### Table S1: PRISMA Checklist for reporting systematic review

| **Section and Topic** | **Item #** | **Checklist item** | **Page where item is reported** |
| --- | --- | --- | --- |
| **TITLE** | | |  |
| Title | 1 | Identify the report as a systematic review. | 1 |
| **ABSTRACT** | | |  |
| Abstract | 2 | See the PRISMA 2020 for Abstracts checklist. | 2 |
| **INTRODUCTION** | | |  |
| Rationale | 3 | Describe the rationale for the review in the context of existing knowledge. | 3 |
| Objectives | 4 | Provide an explicit statement of the objective(s) or question(s) the review addresses. | 3 |
| **METHODS** | | |  |
| Eligibility criteria | 5 | Specify the inclusion and exclusion criteria for the review and how studies were grouped for the syntheses. | 4 |
| Information sources | 6 | Specify all databases, registers, websites, organisations, reference lists and other sources searched or consulted to identify studies. Specify the date when each source was last searched or consulted. | 4-5 |
| Search strategy | 7 | Present the full search strategies for all databases, registers and websites, including any filters and limits used. | 4-5, Table S2 |
| Selection process | 8 | Specify the methods used to decide whether a study met the inclusion criteria of the review, including how many reviewers screened each record and each report retrieved, whether they worked independently, and if applicable, details of automation tools used in the process. | 4-5 |
| Data collection process | 9 | Specify the methods used to collect data from reports, including how many reviewers collected data from each report, whether they worked independently, any processes for obtaining or confirming data from study investigators, and if applicable, details of automation tools used in the process. | 5, Table S3 |
| Data items | 10a | List and define all outcomes for which data were sought. Specify whether all results that were compatible with each outcome domain in each study were sought (e.g. for all measures, time points, analyses), and if not, the methods used to decide which results to collect. | 5, Table S3 |
|  | 10b | List and define all other variables for which data were sought (e.g. participant and intervention characteristics, funding sources). Describe any assumptions made about any missing or unclear information. | 5, Table S3 |
| Study risk of bias assessment | 11 | Specify the methods used to assess risk of bias in the included studies, including details of the tool(s) used, how many reviewers assessed each study and whether they worked independently, and if applicable, details of automation tools used in the process. | 5 |
| Effect measures | 12 | Specify for each outcome the effect measure(s) (e.g. risk ratio, mean difference) used in the synthesis or presentation of results. | n/a |
| Synthesis methods | 13a | Describe the processes used to decide which studies were eligible for each synthesis (e.g. tabulating the study intervention characteristics and comparing against the planned groups for each synthesis (item #5)). | 5-6 |
|  | 13b | Describe any methods required to prepare the data for presentation or synthesis, such as handling of missing summary statistics, or data conversions. | n/a |
|  | 13c | Describe any methods used to tabulate or visually display results of individual studies and syntheses. | 5-6 |
|  | 13d | Describe any methods used to synthesize results and provide a rationale for the choice(s). If meta-analysis was performed, describe the model(s), method(s) to identify the presence and extent of statistical heterogeneity, and software package(s) used. | 5-6 |
|  | 13e | Describe any methods used to explore possible causes of heterogeneity among study results (e.g. subgroup analysis, meta-regression). | n/a |
|  | 13f | Describe any sensitivity analyses conducted to assess robustness of the synthesized results. | n/a |
| Reporting bias assessment | 14 | Describe any methods used to assess risk of bias due to missing results in a synthesis (arising from reporting biases). | n/a |
| Certainty assessment | 15 | Describe any methods used to assess certainty (or confidence) in the body of evidence for an outcome. | n/a |
| **RESULTS** | | |  |
| Study selection | 16a | Describe the results of the search and selection process, from the number of records identified in the search to the number of studies included in the review, ideally using a flow diagram. | 7, Figure S1 |
|  | 16b | Cite studies that might appear to meet the inclusion criteria, but which were excluded, and explain why they were excluded. | Figure S1 |
| Study characteristics | 17 | Cite each included study and present its characteristics. | 7, Table S3 |
| Risk of bias in studies | 18 | Present assessments of risk of bias for each included study. | 7, Table S4 |
| Results of individual studies | 19 | For all outcomes, present, for each study: (a) summary statistics for each group (where appropriate) and (b) an effect estimate and its precision (e.g. confidence/credible interval), ideally using structured tables or plots. | 7, Table S3 |
| Results of syntheses | 20a | For each synthesis, briefly summarise the characteristics and risk of bias among contributing studies. | Table S3 |
|  | 20b | Present results of all statistical syntheses conducted. If meta-analysis was done, present for each the summary estimate and its precision (e.g. confidence/credible interval) and measures of statistical heterogeneity. If comparing groups, describe the direction of the effect. | 8-17 |
|  | 20c | Present results of all investigations of possible causes of heterogeneity among study results. | n/a |
|  | 20d | Present results of all sensitivity analyses conducted to assess the robustness of the synthesized results. | n/a |
| Reporting biases | 21 | Present assessments of risk of bias due to missing results (arising from reporting biases) for each synthesis assessed. | 7, n/a |
| Certainty of evidence | 22 | Present assessments of certainty (or confidence) in the body of evidence for each outcome assessed. | 7, n/a |
| **DISCUSSION** | | |  |
| Discussion | 23a | Provide a general interpretation of the results in the context of other evidence. | 17-19 |
|  | 23b | Discuss any limitations of the evidence included in the review. | 20 |
|  | 23c | Discuss any limitations of the review processes used. | 20 |
|  | 23d | Discuss implications of the results for practice, policy, and future research. | 20 |
| **OTHER INFORMATION** | | |  |
| Registration and protocol | 24a | Provide registration information for the review, including register name and registration number, or state that the review was not registered. | 4 |
|  | 24b | Indicate where the review protocol can be accessed, or state that a protocol was not prepared. | n/a |
|  | 24c | Describe and explain any amendments to information provided at registration or in the protocol. | n/a |
| Support | 25 | Describe sources of financial or non-financial support for the review, and the role of the funders or sponsors in the review. | 21 |
| Competing interests | 26 | Declare any competing interests of review authors. | 21 |
| Availability of data, code and other materials | 27 | Report which of the following are publicly available and where they can be found: template data collection forms; data extracted from included studies; data used for all analyses; analytic code; any other materials used in the review. | Supplementary tables |

### Table S2: Search strategy

- **P**opulation: Women and/or birthing people planning pregnancy, pregnant, or postpartum, in an HIC setting (as classified by the World Bank, 2024), and from an underserved community, defined as individuals or groups with one or more social risk factors, which may have resulted in them being systematically excluded or denied full opportunity to participate in economic, social, or civic life. Social risk factors were as expansive as possible, including: young or advanced maternal age; single mothers; low SES; any group identified as a minority within the study setting (e.g., ethnicity or sexual orientation); refugee, or asylum-seeker; facing homelessness, victim/survivor of domestic abuse; living in a deprived area; having a diagnosed mental health condition or learning disability; physical disability or chronic illness; substance abuse; and not speaking the language local to the country in which the healthcare was provided.
- **E**xposure: All routine antenatal and intrapartum care, comprising planned care before and during labour and birth, to optimise outcomes for mothers and babies, as defined by WHO guidelines (WHO, 2016). This care includes the minimal number of planned antenatal care appointments, health promotion activities (such as advice on healthy diet and exercise), urine and blood tests (such as to screen for anaemia), vaccination and supplementation (such as with iron), monitoring of fetal wellbeing, additional care for women and/or birthing people at higher risk, and care provided for labour and birth (such as for progress in labour and skilled birth attendance). The setting could be within hospitals, the community, or at home.
- **O**utcome: Care-seeking experiences, including health knowledge, behaviours, perceptions, and healthcare utilisation.

| **Review question: Which factors influence the candidacy to seek care during pregnancy of pregnant women and birthing people from under-served groups in high income settings?** | | | | | | | | | |
| --- | --- | --- | --- | --- | --- | --- | --- | --- | --- |
| **Search combination and results** | **Keyword 1**  **(Population)** |  | **Keyword 2 (Population)** |  | **Keyword 3 (Population)** |  | **Keyword 4 (Exposure)** |  | **Keyword 5 (Outcome)** |
|  | High income countries | **AND** | Childbearing women and birthing people | **AND** | Under served groups | **AND** | Maternity services | **AND** | Care-seeking behaviour |
| **Database** | **Search term keyword 1** |  | **Search term keyword 2** |  | **Search term keyword 3** |  | **Search term keyword 4** |  | **Search term keyword 5** |
|  | High-Income OR High-Income Countr* OR High Income OR High Income Countr* OR  HIC OR Andorra OR Greece OR Palau OR (Antigua and Barbuda) OR Greenland OR Panama OR Aruba OR Guam OR Poland OR Australia OR Hong Kong OR Portugal OR Austria or Hungary or Puerto Rico or Bahamas or Iceland or Romania or Bahrain or Ireland or Qatar or Barbados or Isle of Man or San Marino or Belgium or Israel or Saudi Arabia or Bermuda or Italy or Seychelles or British Virgin Islands or Japan or Singapore or Brunei Darussalam or Republic of Korea or South Korea or Saint Maarten or Canada or Kuwait or Slovakia or Cayman Islands or Latvia or Slovenia or Channel Islands or Liechtenstein or Spain or Chile or Lithuania or (Saint Kitts and Nevis) or Croatia or Luxembourg or Saint Martin or Curacao or Macao or Sweden or Cyprus or Malta or Switzerland or Czech Republic or Mauritius or Taiwan or Denmark or Monaco or (Trinidad and Tobago) or Estonia or Nauru or (Turks and Caicos) or Faroe Islands or Netherlands or United Arab Emirates or Finland or New Caledonia or United Kingdom or UK or France or New Zealand or United States or USA or French Polynesia or Northern Mariana Island or Uruguay or Germany or Norway or Virgin Islands or Gibraltar or Oman |  | Childbearing OR Wom?n OR Female OR Pregnant OR Pregnan* OR Birthing People |  | teenage parent* OR young mother* OR young mum* OR socioeconomic factor* OR social class OR equity OR inequalit* OR equalit* OR unequal*OR inequitabl* OR inequit* OR disparit*OR gap* OR disadvantage*OR socioeconomic* OR socioeconomic status OR vulnerable population* OR minority health OR minority group* OR multiethnic* OR multiracial OR multi racial OR refugee* OR immigrant OR migrant* OR asylum seeker* OR traveller OR gyps* OR roma OR roman* OR homeless OR homelessness OR domestic abuse OR domestic violence OR deprived area* OR innercity OR inner city OR innercities OR inner cities OR mental disorders OR mental illness OR mental health problem* OR disabled OR differently abled OR learning disorders OR drug abuse OR substance abuse OR social exclusion OR socially excluded OR social inclusion OR social complexity OR socially complex OR non English  OR communication barriers OR multilingual OR vulnerable populations OR sensitive populations |  | antenatal care OR antenatal clinic* OR antenatal health service* OR antenatal healthcare OR antenatal program* OR antenatal programme* OR antenatal service* OR maternity care OR maternity clinic* OR maternity health service* OR maternity healthcare OR maternity program* OR maternity programme* OR maternity service* OR obstetric care OR obstetric clinic* OR obstetric health service* OR obstetric healthcare OR obstetric program* OR obstetric programme* OR obstetric service* OR perinatal care OR perinatal clinic* OR perinatal health service* OR perinatal healthcare OR perinatal program* OR perinatal programme* OR perinatal service* OR prenatal care OR prenatal clinic* OR prenatal health service* OR prenatal healthcare OR prenatal program* OR prenatal programme* OR prenatal service* OR maternal healthcare OR maternal health service OR prenatal care OR perinatal care OR obstetric nursing OR midwifery OR nurse midwives OR maternal child nursing OR maternal health services OR midwife OR midwifery OR midwives OR vaccination or vaccines or immunisation |  | Health care seeking behavio?r OR care seeking behavio?r OR care-seeking behavio?r OR health care behavio?r OR health behvaio?r OR attitude to health OR care seeking practices OR care seeking patterns OR utilisation OR health service utilisation OR health knowledge OR health practice |

### Table S3: Details of included studies

| **Study ref** | **Country** | **COVID-19 Impact** | **Aim** | **Participant description** | **Data collection** | **Data analysis** | **Summary of reported results** | **Inter-sectional approach** | **Social risk factors** |
| --- | --- | --- | --- | --- | --- | --- | --- | --- | --- |
| Alanazy et al., Midwifery, 2019. | Saudi Arabia | No | To understand the beliefs of pregnant women and health professionals about the factors leading to low attendance rates in ANC | n=21 pregnant women >18 years, who had missed one or more ANC appointments + n=9 HCPs working with new mothers | IDIs | Thematic analysis | Maternal attitudes and beliefs about the importance of care- ANC considered to be unnecessary by some and only to be attended for emergencies, value of ultrasound, maternal attitude influenced by others such as husband, accessibility of care and family duties, transport issues, employment commitments, other child responsibility, facilities at ANC clinics not up to standard, long waiting times for waste of appointment, lack of specialised facilities in certain clinics, preference for private care if they can afford, impact of staff attitudes and knowledge- pressuring behaviour, dismissive staff, poor care- inconsistent advice, misdiagnosis | No | Delayed antenatal care users |
| Ayers et al., Maternal and Child Health Journal, 2018. | USA | No | Examine Marshallese mothers’ beliefs, perceptions, and experiences of prenatal care and to identify potential barriers. | n= 43 mothers, who self-reported as Marshallese, and were 18 years or older and living in Arkansas | Focus Groups | Thematic analysis | Structural barriers include negotiating health insurance, transportation, and language barriers. The social–cultural barriers that emerged included a lack of understanding of the importance of seeking early and consistent prenatal care, as well as how to navigate the healthcare process. | No | Ethnic minority |
| Bains et al., BMC Pregnancy and Childbirth, 2021. | Norway | No | Identify challenges and barriers recently migrated women face in accessing and utilising maternity healthcare services. | N=20 migrant women in urban Oslo, with a length of stay ≤ 5 years in Norway and born in a low- or middle-income country. 7 in-depth interviews with midwives working in Oslo. Findings triangulated with 401 face-to-face questionnaires postpartum at hospitals among migrant women. | Structured questionnaire, IDIs | Thematic analysis | 1) Navigating the healthcare system, (2) Language, (3) Psychosocial and structural factors, and (4) Expectations of care. | No | Migrants, refugees and asylum seekers |
| [Balaam et al., Women and Birth, 2018.](https://pubmed.ncbi.nlm.nih.gov/29370993/) | UK | No | To explore vulnerable/marginalised women's views and experiences of receiving targeted support from a specialist midwifery service and/or a charity. | N-11 Women who had received targeted support | IDIs | Thematic analysis | Enabling needs-led care and support – assures best access to care (co-planning), Designed to fit individual lifestyles, home visits made available, frequency of support, advocacy and other forms of emotional support offered to women during care. Empowering through knowledge – preparation for childbirth through clear explanations, information relevant to particular needs, specialist information. Value of a supportive presence -positive emotional impact of accessing support. Developing capabilities, motivation and confidence (increasing motivation and capabilities to abstain from illegal substances | No | Referred for safeguarding/ child protection concerns |
| Barkensjo et al., BMC Pregnancy and Childbirth, 2018. | Sweden | No | Provide a composite description of women’s experiences of clinical encounters throughout pregnancy and childbirth, when living as undocumented migrants in Sweden. | N=13 Women from 18-36 years of age Macedonia, Romania, Bosnia, Albania, Somalia, Afghanistan, Serbia, Chechnya, Morocco, and Kosovo | Unstructured IDIs | Qualitative content analysis | In clinical encounters where healthcare professionals displayed empathic concern and listening behaviours, women felt empowered, acknowledged, and encouraged, leading them to trust clinicians, diminishing fears relating to seeking healthcare services. Conversely, when neglectful behaviour on part of healthcare professionals was perceived in encounters, anxiousness and fear intensified. | No | Migrants, refugees and asylum seekers |
| [Barkin et al., Journal or Midwifery and Women’s Health](https://onlinelibrary.wiley.com/doi/10.1111/jmwh.13204) | USA | No | To determine the potential acceptability of home visiting services within the vulnerable population and identify what services women want | n=32 women >18 years, English speaking, currently speaking and attending high risk obstetric clinic | Focus groups | Not clearly stated | (1) Realities of high risk pregnancies and need for additional support- appreciated support , (2) Level of current exposure to home visiting- found it helpful with taking care of child, attending to house chores, or taking time for self, (3) Receptive to home visiting- mixed feelings, some appreciated the help others didn’t want strangers in their house or handling their baby, (4) Apprehensive of home visiting- concern of HCP criminal past, judgement over household or life | No | Obstetric high-risk classification |
| Bitar et al., Midwifery, 2020. | Sweden | No | The purpose of this study was to explore Arabic-speaking women’s´ experiences of communication at antenatal care in Sweden when using a tablet application (app) | N= 10 born in an Arabic country, spoke Arabic and now living in Sweden, despite the level of Swedish language skills and had used the App at least two times, different gestational weeks or at postpartum check-up, both primiparous and multiparous. | Telephone interviews | Content analysis | 4 main themes: 1. Adapting the content to the patient group 2. Language and communication, 3. User friendly 4. Improvement proposal | No | Migrants, refugees and asylum seekers |
| [Byatt et al., Obstetrics & Gynaecology, 2018.](https://journals.sagepub.com/doi/10.1177/2150132720944074) | USA | No | To assess utilisation outcomes of MCPAP for Moms program and elicit patient and obstetric care provider perspectives on barriers and facilitators to addressing perinatal mental health and substance use disorders. | n=10 women aged 18-55, English speaking, > 4 weeks gestation- 4 months postpartum, receiving care from a practice enrolled in MCPAP for Moms, EPDS>10, did not meet criteria for active acute mental health disorder | IDIs | Modified grounded theory approach | Perceptions about mental health hindered ability to ask/participate in treatment- stigma, unfit mother judgement, belief that mental health care is outside Obstetric care, signposted to resources but not referred, felt that physical health was prioritised more, EPDS allowed them to inquire about their mental health score and status, normalisation of depression, trusting relationship | No | Mental health problems; substance abuse |
| [Daoud et al., Social Science and Medicine, 2022.](https://www.sciencedirect.com/science/article/pii/S0277953621008832?via%3Dihub) | Israel | No | To use critical race theory and intersectionality to uncover mechanisms of racial maternal separation (RMS) and inequitable maternal care in hospital maternity wards at three levels- policy, practice, and women's experiences | n=26 women who gave birth in study sites: public hospital in three areas of the country + HCPs and head of departments | IDIs | thematic analysis | (1) Mothers who experience RMS- unsure of the reason why, accept over time, some unaware of the concept but report experiencing it; (2) Discrimination based on RMS- Mostly Palestinian Arab or Ultra-orthodox Jewish who experienced it- visibly minority, but RMS felt comfortable to be with other similar women, easier option; (3) Requests for separation did not come from me- no minority women requested, those of higher status/SES/Israeli requested, sometimes under the guise of peace and quiet, but ethno-national motivated- "Arabs tend to have a lot of visitors and is loud", many internalised that it’s a good thing- share the same culture its good for us | Yes | Ethnic and religious minority |
| Decker et al., BMC Pregnancy and Childbirth, 2021. | Mexico, USA | No | Compare the maternity care experiences of Mexican-origin adolescents in Guanajuato, Mexico and Fresno, California from both youth and healthcare provider perspectives | n=74 young women (+15 Healthcare providers) - Female youth were eligible if they were either pregnant or post-partum (within 12 months of delivery), spoke English or Spanish, and were 14–20 years of age. Youth respondents in California were required to either have migrated from Mexico or have at least one parent who had | IDIs, focus groups | Grounded theory | 1.need for communication and clear explanations, 2. respectful versus judgmental providers 3. engaging youth in decision-making 4. focus on the age of the youth and their partners. | No | Migrants, refugees and asylum seekers |
| [Drago et al., Maternal and Child Health Journal, 2018.](https://link.springer.com/article/10.1007/s10995-018-2471-8) | USA | No | To characterise Latino parental perceptions of antenatal counselling in order to validate a Spanish decision-aid to improve parental knowledge of prematurity after antenatal consults. | n=22 Latino parents of infants born before 26 gestational age from 5 participating high-risk follow-up clinics at academic centres in Philadelphia, PA, Evanston, IL, and New York, NY | IDIs | thematic analysis | (1)    Information, (2) Framing of information, (3) Transition to NICU, (4) Factors impeding processing ability, (5) Coping mechanisms, (6) Parental perceptions, (7) Intercultural linguistic barriers | No | Limited local language ability |
| [Farewell et al., Journal or Primary Care & Community Health, 2020.](https://www.sciencedirect.com/science/article/pii/S0266613819301202) | USA | Yes | To use mixed-methods to better understand the mental health and well-being effects of the COVID-19 pandemic, as well as sources of resilience, among women during the perinatal period. | n=21 women, >18 years, English speaking, living in Colorado, and being pregnant or within 6 months postpartum | IDIs | Constant comparative analysis | (1) Uncertainty surrounding care and risk exposure- stress about changes and unknowns, risk of COVID, lack of clear guidance and recommendations; (2) Lack of anticipated support networks and loneliness- lack of social support, less excitement about pregnancy, concern over postpartum support; (3) Factors that support positive coping and resilience- partners primary support, being outdoors and Zoom classes with new moms; (4) Positive impacts of COVID-19 pandemic on perinatal mental health and well-being- more time to prioritise self and self-care, increased connection and bonding with baby and immediate family unit | No | Mental health problems |
| [Frederiksen et al., Journal of Clinical Nursing, 2021.](https://onlinelibrary.wiley.com/doi/10.1111/jocn.15778) | Denmark | No | To identify the key elements of supportive care practices by exploring how parents in vulnerable positions experiences their relationship and encounters with HCPs in maternity care | n=26 women (and their n=13 partners in dyads), received specialist services due to their vulnerability, Danish speaking | IDIs | Thematic analysis | (1) Having a voice/feeling listened to- safe space to voice and discuss their experiences, attentive listening, open questions made it safer to engage, (2) Being met with empathy/feeling understood- felt supported by professionals showing empathy and acknowledging the difficulty of their situation, accommodating specific needs made it easier to share, (3) Worthy of attention/being taken seriously-concerns taken into consideration, acknowledgement of worries, heard and respected, (4) On equal terms- not being judged by scored perception of vulnerability, (5) Moving in the right direction/feeling reassured- HCP positive appraisal and encouragement help relieve parents' self-doubts and concerns, transparency in tackling worries (removal of child and social services) | No | Complex social, medical, or psychological challenges that may require possible involvement of social services; substance abuse, severe mental illness, medical complexity. |
| Frederiksen et al., Qualitative Health Research, 2021 | Denmark | No | To identify what parents fear, how and why they experience it, and how this shapes the childbearing experience and engagement with services | n=26 women (and their n=13 partners in dyads), received specialist services due to their vulnerability, Danish speaking | IDIs | Thematic analysis | (1) Not wanting to go back to that dark place- scared of falling back into mental health struggles and seeking extra support to avoid it, (2) Knowing that I will have an impact on the baby- fear of being a bad mother and falling short, (3) Being labelled as the ones who couldn't - worried about how HCPs would respond to information shared, reported etc based on previous negative experiences, (4) What are they going to set in motion- afraid of HCPs due to their role in child surveillance and removal, consequences of being honest | No | Complex social, medical, or psychological challenges that may require possible involvement of social services; substance abuse, severe mental illness, medical complexity. |
| Gong et al, Midwifery, 2022 | UK | No | Investigate how migrant pregnant women engage with digital tools technologies and resources | N=30;17 Chinese migrant women & 13 midwives | IDIs | Framework analysis | Positive about use of digital resources, concerns related to digital poverty, low eHealth literacy among migrant women. English as language barrier | No | Migrants, refugees and asylum seekers |
| [Goodwin et al., Health Expectations, 2018.](https://www.ncbi.nlm.nih.gov/pmc/articles/PMC5750740/pdf/HEX-21-347.pdf) | UK | No | Exploring the importance of midwife–woman relationships in care satisfaction and pregnancy outcomes for migrant and minority ethnic women in the UK | 9 Migrant Pakistani participants 11 practising midwives | Ethnography | Thematic analysis | Family relationships are huge influence, seen as a barrier by midwives. Culture/religion also embedded throughout pregnancy (traditions and postnatal practices) differ from UK system | No | Migrants, refugees and asylum seekers |
| [Gordon et al., British Journal of General Practice, 2019.](https://pubmed.ncbi.nlm.nih.gov/31501164/) | UK | No | Explore perspectives of women who have experienced pregnancy and homelessness to ascertain how to improve perinatal care | n=11 women from 3 facilities for individuals experiencing homelessness (shelter, short term residence, and day-care facility) | IDIs | Iterative thematic analysis | Unstable family and childhood trauma – inadequate health knowledge - perceived norms and an expectation of stigma because of extensive adversity and low self-esteem produced a mistrust of health and social care practitioners. Wanting the best for the baby versus fear of child loss- 1st time women felt dependent on -motivation to attend apps – mistrust meant women did not receive professional support. Biomedically competent, emotionally unsupportive care. | No | Homelessness |
| Hall et al., BMC Pregnancy and Childbirth, 2018. | UK | No | Explore the experiences of dignity and respect in childbirth of women with disability. | N= 37 women | Survey with open-ended question | Thematic analysis | Positive responses of care received- information and services. Dissatisfied with- individuality and preferences were respected. Maternity care- care providers awareness and attention to main impact of disabilities (midwife on day of delivery not knowing/no notes read, labour ward unaware), Choice, Dignity and respect. | No | Physical, sensory, and mobility impairment or disability |
| [Henry et al., Qualitative Health Research, 2019.](https://journals.sagepub.com/doi/10.1177/1049732319873620) | Germany | No | To investigate how premigration experiences, conceptions about pregnancy and childbirth, health literacy and language skills influence access to health care, experiences of health care, and childbirth. | n=12 women >18 years, having an ongoing or completed asylum procedure in Germany, being pregnant or having given birth to a child in Germany | IDIs | Content Analysis | (1) Concepts of pregnancy and childbirth, premigration experiences- closely related to beliefs of healthcare from home, information passed on by female relatives, only visiting for ultrasound and birth, no knowledge about non-invasive pain relief, CS considered helpful, (2) Impact of premigration experiences on the women's perceptions of health care needs- used to continuity of care in home country, vaginal exam/limitation in number of US scans caused anxiety as not common elsewhere, ANC services partially or entirely unknown as not offered in home country, lack of single contact point left women not knowing when to go to hospital/waiting too long, (3) Experiences of antenatal and obstetric care and compensation mechanism for access barriers- refugee clinic easily accessible as did not need to make appointment, care was culturally appropriate (female OBGYN with interpreter) but wait times long, other clinics quicker but asked to bring own interpreters which was difficult, care administered without patient fully understanding- feeling of powerlessness, language barriers and knowledge deficits, resorted to Arabic information online | No | Limited local language ability; Migrants, refugees and asylum seekers |
| Hjelm et al., BMC Women’s Health, 2018. | Sweden | No | To explore development over time, during and after pregnancy, of beliefs about health, illness and healthcare in migrant women with GDM born in Africa living in Sweden, and study the influence on self-care and care seeking | N=9 Women, (23–40 years), on three different occasions: during pregnancy (gestational weeks 34–38), and 3 and 14 months after delivery managed at an in-hospital diabetes specialist clinic in Sweden | IDIs | Content analysis | 1. Beliefs about illness (both prior and post diagnosis) 2. Beliefs about health (how illness impacted their help postpartum) 3. Beliefs about healthcare (experience of the system during diagnosis and pregnancy) | No | Migrants, refugees and asylum seekers; medical complexity |
| Holden et al., Asia-Pacific Psychiatry, 2019. | New Zealand | No | Explored current maternal mental health (MH) screening practices and supports | 13 mothers participated (Tongann=7,Maorin=5, and New Zealand European= 1, Mothers participating in a parenting programme designed for new mothers | IDIs, focus groups | Thematic analysis | 1.Screening is ad-hoc. 2.MH supports are variable and inconsistent. 3.Barriers hamper screening and access to supports. 4.Service improvements should be multileveled. | No | Indigenous groups |
| Holten et al., Qualitative Health Research, 2018. | The Netherlands | No | Defining moments for Women Choosing Home Birth in High-Risk Pregnancies in The Netherlands | 10 cases in which Dutch women with a high-risk pregnancy chose to birth at home against medical advice | IDIs | Theoretical Framework | Previous trauma, weighing in evidence, Paternalistic decision making and a perceived lack of autonomy, Inflexibility: Conflict in negotiation, Holistic care: A last resort/second best choice, Defining moment. | No | Decision to birth outside the system |
| [Husain et al., PLoS One, 2022.](https://www.ncbi.nlm.nih.gov/pmc/articles/PMC9385003/) | UK | Yes | Explore COVID-19 vaccination uptake, facilitators and barriers in ethnically-diverse pregnant women. | N=441 Pregnant women attending routine antenatal clinic appointments | Survey with open-ended question | Thematic analysis | Vaccine hesitancy- Concerns regarding safety, effects of vaccine. Lack of trust in both vaccines and UK system, low participation in research from ethic minority group (personal/structural barriers). | No | SES; ethnic minority |
| Ikhilor et al., Journal of Advanced Nursing, 2019. | Switzerland | No | To describe communication barriers faced by allophone migrant women in maternity care provision from the perspectives of migrant women, healthcare professionals, and intercultural interpreters | N= 36, four Albanian and six Tigrinya speaking women, 22 healthcare professionals and four intercultural interpreters | IDIs | Thematic analysis | 1.The challenge of understanding each other's world 2. Communication breakdowns 3. Imposed health services. | No | Limited local language ability |
| Johnsen et al., International Journal of Environmental Research and Public Health, 2020. | Denmark | No | The study examines the context of the implementation of the MAMAACT intervention and investigates how the intended intervention mechanisms regarding response to pregnancy complications were affected by barriers in non-Western immigrant women’s everyday life situations. | N=21 non-Western immigrant women who received the MAMAACT leaflet, being at least 28 weeks pregnant, and being at least 18 years old | IDIs | Systematic text condensation & situational-adaptation framework | 1.Sources of knowledge during pregnancy 2. Containment of pregnancy warning signs 3. Barriers during the onset of acute illness 4.Previous situations with maternity care providers | No | Migrants, refugees and asylum seekers |
| [Kuipers et al., Women and Birth, 2022.](https://www.sciencedirect.com/science/article/pii/S1871519221000391?via%3Dihub) | The Netherlands | No | To examine if pregnant migrant women make sense of vulnerability during pregnancy | n= 25 pregnant women, >16 years, with Western and non-Western migrant background, speaking Dutch, given birth in Dutch system since 2009. | Focus groups | Thematic analysis | (1) Look beyond who you think I am and see and treat me for who I really am- right to be human, not regarded as a vulnerable category based on a screening questionnaire, (2) Ownership of truth and knowledge- HCPs have the need to control their health and birth outcomes- wish to be less authoritative and ignorant; did not think they were more vulnerable that Dutch women living in similar and more affluent areas, (3) Don't punish me for being honest-experienced or witnessed CPS removing children, felt punished, consequences with partner in case of DV- distrust, (4) Projection of fear- HCPs instilling fear in mothers based on vulnerability, focused on clinical outcomes rather than social and emotional aspects, (5) Coping with labelling- delayed care to stay in control and regain humanity lost by labelling them as vulnerable (aware that this leads to poor outcomes). | No | Migrants, refugees and asylum seekers |
| [McClellan et al., Journal of Transcultural Nursing, 2022.](https://journals.sagepub.com/doi/10.1177/10436596221091689?url_ver=Z39.88-2003&rfr_id=ori:rid:crossref.org&rfr_dat=cr_pub%20%200pubmed) | USA | No | The purpose of this research is to explore the lived experiences of Mongolian immigrant women seeking perinatal/peripartum care in the United States | n=12 Mongolian-born women currently living in the United States who accessed health care during pregnancy or delivery were eligible for inclusion in this study. | IDIs | Thematic content analysis | (1) Searching for support- support from family especially mothers is culturally important, or alternative sources of support is critical- wanted more support from HCP (with understanding Mongolian cultural practices- didn’t take time to get to know them as a person), (2) Communication and Education- miscommunication even though all spoke good English such as didn’t know birth plan meaning, navigating educational material difficult in second language, (3) Comparing and contrasting the health care systems- differences between home country and navigating new system, grateful for some difference in US system such as fathers being allowed in, (4) Traditions worth keeping- all wanted to keep some traditions, some of which were immediately challenged in USA system. | No | Migrants, refugees and asylum seekers |
| McLeish et al., Women and Birth, 2019. | UK | No | Explore the maternity experiences of mothers each of whom was vulnerable through multiple challenges | n=40 mothers experiencing multiple disadvantages defined as low SES and at least one additional factor: Aged under 25, recent migrants, asylum seekers and refugees, mothers from BAME communities, single parents, living with physical or mental illness and domestic abuse. | IDIs | Thematic analysis | A confusing and frightening experience – complex system, language barrier, lack of information leads to people turning to other sources of information; Longing to be respected as an individual – midwives giving full attention to patients, relationship with doctors; Trust – also negative experience with staff; Importance of choice and control -decisions about maternity care, needing trust to feel safe. | No | Low SES; young mothers; single mothers; migrants, refugees and asylum seekers; ethnic minority; physical or mental illness; domestic abuse |
| McLemore et al., Social Science and Medicine, 2018. | USA | No | Study described the pregnancy-related healthcare experiences of women of colour from Fresno, Oakland, and San Francisco, California, with social and/or medical risk factors for preterm birth. | n=54 Women from Fresno, Oakland, and San Francisco who were age 18 or greater, English speaking, English and/or Spanish Speaking in Fresno, with medical and/or social risk factors for PTB from community-based programs. | Focus groups | Thematic analysis | 1. Disrespect 2. Stressful interaction 3. Inconsistent social support 4. Unmet information needs 5. Perceived competence and confidence in parenting and newborn care. | No | Ethnic minority; medical complexity |
| [Mehrara et al., Empirical Studies, 2022.](https://www.tandfonline.com/doi/full/10.1080/17482631.2022.2066256) | Norway | No | To explore how immigrant women from diverse countries and ethnic backgrounds experienced and navigated the Norwegian maternal health service during pregnancy and childbirth. | n=11, immigrant women who had carried out pregnancy and childbirth in Norway, recruited from public kindergartens and community; contingent purposive sampling. | IDIs | Thematic analysis | Reflection on differences and similarities to home country; 1. Approach to pregnancy: different levels of medicalisation of pregnancy in different countries, independence empowered in Norway (not in home country). 2. Monitoring and follow ups: lack of regular testing in welfare healthcare systems (not as much importance given before birth because postnatal support is available). Not trusting of Norwegian system and staff competence. proactivity in bridging the gap between her expectations of maternal health care, which was not only shaped but continually influenced by medical practice and advice from her social network in her home country. 3. Approach to birth: desire for ELCS not common in Norway, developed confidence for natural birth through trusting relationship with midwife. 4. Communication during labour- communication not language was lacking. Not involved in decisions, treated like a passive participant | No | Migrants, refugees and asylum seekers |
| Mule et al., Women and Birth, 2022. | Australia | No | To assess pregnant women's attitude to and reasons for non-disclosure at, comprehensive psychosocial assessment with their midwife. | n= 161 women (n=1976 total survey responses), who attended first antenatal scan at study site between 28 March 2017- 27 May 2019 | Survey with open-ended question | Thematic analysis | (1) Normalising and negative self-perception- attributing poor mental health to pregnancy hormones, and ashamed to admit they were not coping well, (2) Fear of negative perception from others- fear and worry about being judged, bad mother, (3) Lack of trust in midwife- most common reason not believing midwife could do anything, not willing to share for fear of care being changed, (4) Women's expectations of appointment- appointment should be about baby not themselves or had forgotten/was unprepared for questions about mental health, (5) Mode of assessment and time issue- limited time and close ended format of questions. | No | Mental health problems |
| [Naylor Smith et al., BMC Pregnancy and Childbirth, 2018.](https://pubmed.ncbi.nlm.nih.gov/29669527/) | UK | No | Identify what influences multiparous women’s choice of birthplace, and to explore their views of home birth. | N=28 | Focus Groups | Thematic analysis | Unfamiliar concept of homebirth to women. Potentially seen as impractical or not affordable, those from minority ethnic backgrounds questions practicality of homebirth, lack of availability or willingness of relatives trust and confidence in maternity system (sources of info related to homebirth) role of HCPS in influencing decisions on choice of birthplace. | No | Ethnic minority; SES |
| [Njenga et al., Journal of Transcultural Nursing, 2023.](https://journals.sagepub.com/doi/10.1177/10436596221125893) | USA | No | The purpose of this study was to explore the resettled Somali refugee women’s experiences and perceptions of maternal health care in the United States. | n=15 Somali refugee women, 18 to 42 years old, spoke English or Swahili, resettled in the United States within the last 5 years, and had accessed the USA maternal health care service with at least one childbirth in the United States. | IDIs | Interpretive descriptive analysis | 1. Communication and resource provision: closer relationship with HCPs who spoke their language, polite, attentive, showed interest in her life outside of pregnancy. Follow up calls were appreciated as a sign of concern, lack of interpreters. 2. Participatory decision making: frustration with paternalistic approach, made to feel dumb, not involved in decisions, miscommunication. 3. Provider attitudes cultural practices: negative perception, stigma and judgement surrounding FGC. 4. Understanding the USA healthcare system: different from previous experiences, difficult to navigate. 5. Mistrust of western health care: belief that tests and procedures are pushed on patients for profit, fear and uncertainty from lack of understanding of procedures- helplessness. 6. Religious beliefs- whatever happens will happen because God's plan | No | Migrants, refugees and asylum seekers |
| Nottingham-Jones et al., Midwifery, 2020. | Australia | No | To explore the lived experiences of preparing for childbirth for nulliparous women aged 35–44 and determine how mature first-time mothers’ can be better supported regarding childbirth preparation. | n=14 nulliparous women aged 35–44 were recruited in their third trimester of pregnancy. | IDIs | Phenomenological Analysis | 1.Building confidence for childbirth 2. Sharing experiences 3. Independent research and preparation 4. Agency and informed decision making 5. Managing the balance between birth planning and flexibility | No | Advanced maternal age |
| Oza-Frank et al., Maternal and Child Health Journal, 2018. | USA | No | To describe the healthcare experiences of a diverse, low-income sample of women with prior GDM, including their suggestions for improving care | N=12 African American, Hispanic, or Appalachian women, aged 18–45 years, and with GDM diagnosis within the past 10 years | Focus Groups | Thematic analysis | Barriers to GDM care, management and follow up - 1. Communication issues 2. Personal and environmental barriers 3. Type and quality of healthcare | No | Ethnic minority; rural setting; medical complexity |
| [Reid et al., Birth, 2021.](https://onlinelibrary.wiley.com/doi/10.1111/birt.12551) | USA | No | To identify barriers and facilitators to early prenatal care of women in Florida | n= 55, women who delayed ANC more than 14 weeks or received none at all | IDIs | Not clearly stated | (1) Personal factors- awareness of pregnancy, mental health, considering abortion, other life factors, (2) Community/social conditions- transportation, social support, (3) Health care system- delay at the clinic level (because of not accepting Medicaid, too high risk), cost of care/insurance, fear or stigma and discrimination, language barriers | No | Delayed antenatal care users |
| Sami et al., Reproductive Health, 2019. | Switzerland | No | Explore positive and negative experiences with maternal health services in the University Hospitals of Geneva and Zurich and to describe barriers to maternity services from a qualitative perspective. | N= 33 women aged 21 to 40 years-with migrant women who were either pregnant or in their first year after childbirth in Geneva and Zurich. | Focus groups | Thematic analysis | Positive results=availability of maternity services, especially during emergency situations and the postpartum period, but also the availability of specific maternity services for undocumented migrants in Geneva. Negative results =personal and structural barriers | No | Migrants, refugees and asylum seekers |
| [Savory et al., Midwifery, 2022.](https://www.sciencedirect.com/science/article/pii/S0266613822000286?via%3Dihub) | UK | No | To explore the experiences of women during pregnancy with mild to moderate mental health problems and describe the barriers to receiving support in relation to their mental health. | n=20 women >18 years, English language, and a viable pregnancy of <18 weeks gestation confirmed by ultrasound scan | IDIs | Thematic analysis | (1) Moods and emotions- past, present and future- mental health over life, past trauma, anxiety- scared of how it will be during pregnancy with heightened hormones; (2) Expectations and control- comparing to others, feeling like a bad mum, searching for information to be in control (online not HCPs); (3) Knowledge and conversations- few had looked for help with mental health, normalising issues, fear of stigma in sharing with others | No | Mental health problems |
| Spangaro et al., Violence against Women, 2019. | Australia | No | Explore evidence for pathways to impact | N=32 abused women; English speaking 28+ weeks pregnant. All participants had experienced past 12 months or current IPV or fear of a partner at the time of screening | IDIs | Qualitative comparative analysis | Key conditions for positive impact 1. Care in the way the midwife asked about the abuse and Support and validation in response to disclosing. Different pathways to positive impact reflect the women's diverse experiences of prior service use, relationship status, contact with statutory agencies and execution of screening. Nil impact= importance of support and validation - absence of those led to negative impact. | No | IPV |
| [Stacey et al., BMC Pregnancy and Childbirth, 2021.](https://bmcpregnancychildbirth.biomedcentral.com/articles/10.1186/s12884-021-03879-2) | UK | No | Explore migrant women’s awareness of health messages to reduce stillbirth risk, and how key public health messages can be made more accessible. | N=30 Migrant women, 18 countries and across 4 National Health Service (NHS) hospitals | Semi-structured focus groups, IDIs | Thematic analysis | Lack of awareness around stillbirth, role of health professionals in terms of understanding structural and cultural barriers during pregnancy. Importance of health messages to keep baby safe during pregnancy. | No | Migrants, refugees and asylum seekers |
| Tesfalul et al., Patient Education and Counselling, 2021. | USA | No | To describe and compare how obstetric patients and care providers view preterm birth risk assessment and communication. | N= 35 18+ speaking English/Spanish | Focus Groups, IDIs | Grounded theory | Preparedness as benefit to preterm birth for patients while providers cited maternal blame, patient alienation, and estimate uncertainty as potential risks. | No | Medical complexity |
| Utne et al., Midwifery, 2020. | Norway | No | To explore Somali women's experiences of antenatal care in Norway. | n=8 Somali-born (first generation) women living in Norway. 22 to 35, | IDIs | Systematic text condensation (STC); a thematic cross-case analysis | 1) when care was provided in a way that gained their trust, they made better use of the available health services, 2) the importance of continuity of care and of sharing commonalities with the caregiver, 3) a need for accessible information, specifically tailored to the needs of Somali women and 4) how culturally insensitive caregivers had a negative impact on the quality of care. | No | Migrants, refugees and asylum seekers |
| Vang et al., Qualitative Health Research, 2018. | Canada | No | We examine patient–provider interactions for Indigenous childbirth evacuees | n=25 Inuit and First Nations women with medically high-risk pregnancies who were transferred or medevacked from northern Quebec to receive maternity care at a tertiary hospital in a southern city in the province. | IDIs | Grounded theory | 1.Evacuation-Related Stress. 2.Hospital Bureaucracy 3. Stereotypes | No | Indigenous groups |
| [Wah et al., Women and Birth, 2019.](https://www.sciencedirect.com/science/article/pii/S1871519217303244?via%3Dihub) | Australia | No | To explore the understanding of self-management experiences of GDM among Chinese migrants | n=18 migrant women of Chinese ethnicity residing in Australia, English/Mandarin/Cantonese speaking, pregnant with a diagnosis of GDM | IDIs | Thematic analysis | (1) Knowledge of GDM- most had good understanding of the problem, contributing factors, future risk; (2) Self-management issues- difficult to manage with traditional Chinese food, eating out with family, physical activity goals difficult to maintain, did not feel the need to do if glucose levels were under control, difficulty with measurement devices and interpreting results, difficult to measure for women with jobs/other children, fear of insulin, made up values to avoid being prescribed insulin; (3) Social support- from partner and family to help self-management- remind of diet, go for walks, hospital primary source of information along with online resources and peers, majority felt comfortable asking questions of HCPs (some felt rushed); (4) Chinese cultural influences- family oriented values bigger than the individual needs, Chinese food- several things not included in guidance (e.g. lotus root), traditional Chinese medicines not seen to be helpful for short term conditions like GDM | No | Migrants, refugees and asylum seekers; medical complexity |
| [Wilson et al., BMC Public Health, 2019.](https://www.ncbi.nlm.nih.gov/pmc/articles/PMC6434850/) | UK | No | Gain a contextualised understanding of factors influencing vaccination acceptance during pregnancy in Hackney, a borough in north-east London, UK. | 47 pregnant and recently pregnant women | IDIs | Thematic analysis | Access (middle class believed they had too much / overwhelmed with sources of info. English not as a first language is a barrier. Healthcare rhetoric- women viewed as irresponsible and in need of management. Community and family influences on vaccination decisions Healthcare professionals’ views towards maternal vaccination. Patient-healthcare professional relationships important. | No | SES; ethnic minority |
| Results of updated search | | | | | | | | | |
| Faulks et al., WOMBI, 2024 | Australia | No | To explore the barriers and enablers that exist for rural women in Australia in accessing perinatal care. | n=19, had a baby in the last 2–8 months and lived in a rural or remote community in Victoria, Australia | IDIs | Reflexive thematic analysis | 1. An individual’s personal characteristics that impact on their capacity to access perinatal care- financial, employment conditions, transportation and distance to care; rural social capital, advocacy, health literacy. 2. Therapeutic relationships and communication- Ineffective relationships, Poor communication; Accessible care providers, Nurturing, safe relationships. 3. How care is organised and delivered by health care services and care providers- under resourced services, differences between areas; technology enables, access to continuity of care. 4. How care providers, health services and communities collaborate to provide perinatal care services- ineffective or effective collaboration between organisations. 5. Policy, funding and governance models that underpin perinatal care services- resource allocation, centralisation of services | No | SES; rural |
| George et al., Midwifery, 2024. | USA | No | Decision-making experiences of people with Medicaid insurance who chose to give birth in a birth centre (BC) in Massachusetts by gathering interview data to interpret and provide meaning about their selection of birth setting. | n=12, people who had a singleton birth at the BC during the 12 months preceding their scheduled interviews. Eligibility criteria included participants who were aged 18 years or older, able to read and speak English, and had Medicaid for insurance coverage | IDIs | Reflexive thematic analysis | 1) Stepping Away from “the System,” 2) Decision-Making with External Influences, 3) Accessing BC Care, 4) Finding a Home at the BC, and 5) Decision-Making as a Temporal Process | No | SES |
| Nechaeva et al., Global Health Action, 2024. | Russia | No | To explore the general awareness of PPD among both midwives and pregnant women in the Arctic Russian town of Arkhangelsk with the further purpose of creating a preliminary model of a postpartum depression prevention program based on empirical data. | n=12 both primiparous and multiparous women between 28–38 weeks of gestation with an uncomplicated pregnancy, residing in Arkhangelsk and over 18 years of age. | IDIs | Systematic text condensation and Phenomenological analysis | 1. Seen as a mix of psychological and physiological symptoms- feelings of apathy; fear, guilt and shame; Bodily- and hormonal changes; Conflicts between expectations and reality; impaired mother-child relationships. 2. Need for professional and family support- Limited time for psychological counselling, Restricted understanding among family members, Financial and housing worries. 3. A medical diagnosis or just temporary fatigue?- High prevalence of symptoms; Uncertainty about labelling; Denial of the problem | No | Rural; mental health problem |
| Parker et al., Qualitative Health Research, 2025. | New Zealand | No | To understand how intersecting marginalizations experienced by our participants, based on their social identities, amplified their exclusion from perinatal services | n=20; 18 years or older; being trans, non-binary, intersex, or Takatāpui and gender-diverse; and having accessed fertility, pregnancy, or birth care within the past 6 years in Aotearoa | IDIs | Reflexive thematic analysis | 1. Perceiving Intersectional Failure- Not Fitting the Normative Script in Perinatal Care, Occupying Multiple Marginalized Identities Seen as “Too Much” by Care Providers; 2. The gaps widen when trans people experience multiple forms of exclusion- The Cards Are Stacked Against You, Falling Through the Gaps; 3. Employing strategies to navigate intersecting exclusions-Deploying Resources to “Conform” to the Script, Reimagining Intersectional Care | Yes | Transgender; Indigenous groups; young pregnancy; sexual and gender minority groups; ethnic minority groups; disability; mental health problems; SES |
| Pierce et al., Front. Digit. Health., 2023. | UK | No | To investigate the impact of digital health on vulnerable pregnant women | n=22 women registered for antenatal care | Survey with open-text question | Thematic analysis | 1. Positive- Convenience and usefullness; communication with HCP;2. Negative- lack of motivation; limited language options; confusing interface, complex app journey; privacy concern, poor communication; 3. Neutral- preference for non-digital | No | IPV; mental health problems; young pregnancies; previous interaction with social services; substance abuse; learning or physical disability; homelessness; migrants, refugees, and asylum seekers; limited local language ability; criminal records |
| Yuill et al., Br. J. Health Psychol., 2024. | UK | No | To explore women's experiences and views of assessment for perinatal anxiety, with the aim of understanding the extent to which they find this acceptable | n=41 if they were pregnant or within 6 weeks of giving birth, aged 16 or over and with sufficient English language | IDIs | Thematic analysis | 1. Raising awareness and improving support, 2. Surveillance and stratifying care, 3. Personalizing care and building trust | No | Mental health problems |

ANC: antenatal care; IDI; HCP: healthcare professional; RMS: racial maternal separation; SES: socioeconomic status; NICU: neonatal ICU; GDM: gestational diabetes; MH: mental health; BAME: Black, Asian, and Minority ethnicity; PTB: preterm birth; ELCS: elective caesarean section; EPDS: Edinbugh Postnatal Depression Score; IPV: intimate partner violence; NHS: National Health Service (UK)

### Table S4: Quality appraisal of included studies

| Study reference | **1. Was there a clear statement of the aims of the research?** | **2. Is a qualitative methodology appropriate?** | **3. Was the research design appropriate to address the aims of the research?** | **4. . Was the recruitment strategy appropriate to the aims of the research?** | **5. Was the data collected in a way that addressed the research issue?** | **6. Has the relationship between researcher and participants been adequately considered?** | **7. Have ethical issues been taken into consideration?** | **8. Was the data analysis sufficiently rigorous?** | **9. Is there a clear statement of findings?** | **10. How valuable is the research?** | **Total Score** |
| --- | --- | --- | --- | --- | --- | --- | --- | --- | --- | --- | --- |
| Alanazy et al., Midwifery, 2019. | 2 | 2 | 2 | 2 | 2 | 1 | 1 | 2 | 2 | 1 | 17 |
| Ayers et al., Maternal and Child Health Journal, 2018. | 2 | 2 | 2 | 2 | 2 | 1 | 2 | 2 | 2 | 2 | 19 |
| Bains et al., BMC Pregnancy and Childbirth, 2021. | 2 | 2 | 2 | 2 | 2 | 0 | 2 | 2 | 1 | 2 | 17 |
| [Balaam et al., Women and Birth, 2018.](https://pubmed.ncbi.nlm.nih.gov/29370993/) | 2 | 2 | 2 | 2 | 2 | 0 | 1 | 2 | 2 | 2 | 17 |
| Barkensjo et al., BMC Pregnancy and Childbirth, 2018. | 2 | 2 | 2 | 2 | 2 | 1 | 2 | 2 | 0 | 0 | 15 |
| [Barkin et al., Journal or Midwifery and Women’s Health](https://onlinelibrary.wiley.com/doi/10.1111/jmwh.13204) | 2 | 2 | 2 | 2 | 2 | 1 | 1 | 2 | 0 | 2 | 16 |
| Bitar et al., Midwifery, 2020. | 2 | 2 | 2 | 2 | 2 | 2 | 2 | 2 | 2 | 2 | 20 |
| [Byatt et al., Obstetrics & Gynaecology, 2018.](https://journals.sagepub.com/doi/10.1177/2150132720944074) | 2 | 2 | 2 | 2 | 2 | 1 | 1 | 2 | 1 | 1 | 16 |
| [Daoud et al., Social Science and Medicine, 2022.](https://www.sciencedirect.com/science/article/pii/S0277953621008832?via%3Dihub) | 2 | 2 | 2 | 2 | 2 | 1 | 1 | 2 | 2 | 2 | 18 |
| Decker et al., BMC Pregnancy and Childbirth, 2021. | 2 | 2 | 2 | 2 | 2 | 2 | 1 | 2 | 2 | 2 | 19 |
| [Drago et al., Maternal and Child Health Journal, 2018.](https://link.springer.com/article/10.1007/s10995-018-2471-8) | 2 | 2 | 1 | 2 | 2 | 0 | 0 | 2 | 2 | 2 | 15 |
| [Farewell et al., Journal or Primary Care & Community Health, 2020.](https://www.sciencedirect.com/science/article/pii/S0266613819301202) | 2 | 2 | 2 | 2 | 2 | 1 | 2 | 2 | 0 | 2 | 17 |
| [Frederiksen et al., Journal of Clinical Nursing, 2021.](https://onlinelibrary.wiley.com/doi/10.1111/jocn.15778) | 2 | 2 | 2 | 2 | 2 | 1 | 2 | 2 | 2 | 0 | 17 |
| Frederiksen et al., Qualitative Health Research, 2021 | 2 | 2 | 2 | 2 | 2 | 2 | 2 | 2 | 2 | 2 | 20 |
| Gong et al, Midwifery, 2022 | 0 | 2 | 2 | 2 | 2 | 1 | 2 | 2 | 2 | 0 | 15 |
| [Goodwin et al., Health Expectations, 2018.](https://www.ncbi.nlm.nih.gov/pmc/articles/PMC5750740/pdf/HEX-21-347.pdf) | 2 | 2 | 2 | 2 | 2 | 2 | 2 | 2 | 2 | 2 | 20 |
| [Gordon et al., British Journal of General Practice, 2019.](https://pubmed.ncbi.nlm.nih.gov/31501164/) | 2 | 2 | 2 | 2 | 1 | 1 | 1 | 2 | 2 | 2 | 17 |
| Hall et al., BMC Pregnancy and Childbirth, 2018. | 2 | 2 | 2 | 2 | 2 | 2 | 2 | 2 | 2 | 0 | 18 |
| [Henry et al., Qualitative Health Research, 2019.](https://journals.sagepub.com/doi/10.1177/1049732319873620) | 2 | 2 | 2 | 2 | 1 | 1 | 1 | 2 | 2 | 2 | 17 |
| Hjelm et al., BMC Women’s Health, 2018. | 2 | 2 | 2 | 1 | 2 | 1 | 1 | 2 | 2 | 2 | 17 |
| Holden et al., Asia-Pacific Psychiatry, 2019. | 2 | 2 | 2 | 2 | 2 | 1 | 1 | 1 | 2 | 2 | 17 |
| Holten et al., Qualitative Health Research, 2018. | 2 | 2 | 2 | 2 | 2 | 2 | 2 | 2 | 2 | 2 | 20 |
| [Husain et al., PLoS One, 2022.](https://www.ncbi.nlm.nih.gov/pmc/articles/PMC9385003/) | 2 | 2 | 2 | 2 | 2 | 1 | 1 | 2 | 2 | 0 | 16 |
| Ikhilor et al., Journal of Advanced Nursing, 2019. | 2 | 2 | 2 | 2 | 2 | 1 | 2 | 2 | 1 | 2 | 18 |
| Johnsen et al., International Journal of Environmental Research and Public Health, 2020. | 2 | 2 | 2 | 2 | 2 | 2 | 2 | 2 | 0 | 2 | 18 |
| [Kuipers et al., Women and Birth, 2022.](https://www.sciencedirect.com/science/article/pii/S1871519221000391?via%3Dihub) | 2 | 2 | 2 | 2 | 2 | 1 | 2 | 2 | 2 | 0 | 17 |
| [McClellan et al., Journal of Transcultural Nursing, 2022.](https://journals.sagepub.com/doi/10.1177/10436596221091689?url_ver=Z39.88-2003&rfr_id=ori:rid:crossref.org&rfr_dat=cr_pub%20%200pubmed) | 2 | 2 | 2 | 2 | 2 | 2 | 2 | 2 | 1 | 0 | 17 |
| McLeish et al., Women and Birth, 2019. | 2 | 2 | 2 | 2 | 2 | 2 | 1 | 2 | 2 | 0 | 17 |
| McLemore et al., Social Science and Medicine, 2018. | 2 | 2 | 2 | 2 | 2 | 2 | 2 | 2 | 2 | 2 | 20 |
| [Mehrara et al., Empirical Studies, 2022.](https://www.tandfonline.com/doi/full/10.1080/17482631.2022.2066256) | 2 | 2 | 2 | 2 | 2 | 1 | 1 | 2 | 2 | 0 | 16 |
| Mule et al., Women and Birth, 2022. | 2 | 2 | 0 | 1 | 1 | 1 | 2 | 2 | 1 | 2 | 14 |
| [Nancy et al., Obstetrics & Gynaecology, 2018.](https://journals.sagepub.com/doi/10.1177/2150132720944074) | 2 | 2 | 2 | 2 | 2 | 1 | 1 | 2 | 1 | 1 | 16 |
| [Naylor Smith et al., BMC Pregnancy and Childbirth, 2018.](https://pubmed.ncbi.nlm.nih.gov/29669527/) | 2 | 2 | 2 | 2 | 1 | 1 | 2 | 2 | 2 | 2 | 18 |
| [Njenga et al., Journal of Transcultural Nursing, 2023.](https://journals.sagepub.com/doi/10.1177/10436596221125893) | 2 | 2 | 2 | 2 | 2 | 2 | 2 | 2 | 2 | 2 | 20 |
| Nottingham-Jones et al., Midwifery, 2020. | 2 | 2 | 2 | 2 | 2 | 1 | 2 | 2 | 1 | 1 | 17 |
| Oza-Frank et al., Maternal and Child Health Journal, 2018. | 2 | 2 | 2 | 2 | 2 | 1 | 1 | 2 | 2 | 2 | 18 |
| [Reid et al., Birth, 2021.](https://onlinelibrary.wiley.com/doi/10.1111/birt.12551) | 2 | 2 | 2 | 2 | 2 | 1 | 1 | 2 | 2 | 2 | 18 |
| Sami et al., Reproductive Health, 2019. | 2 | 2 | 2 | 2 | 2 | 1 | 1 | 2 | 2 | 2 | 18 |
| [Savory et al., Midwifery, 2022.](https://www.sciencedirect.com/science/article/pii/S0266613822000286?via%3Dihub) | 2 | 2 | 2 | 2 | 2 | 1 | 2 | 2 | 2 | 2 | 19 |
| Spangaro et al., Violence against Women, 2019. | 2 | 2 | 2 | 2 | 2 | 1 | 2 | 2 | 2 | 2 | 19 |
| [Stacey et al., BMC Pregnancy and Childbirth, 2021.](https://bmcpregnancychildbirth.biomedcentral.com/articles/10.1186/s12884-021-03879-2) | 2 | 2 | 2 | 1 | 2 | 0 | 2 | 2 | 2 | 2 | 17 |
| Tesfalul et al., Patient Education and Counselling, 2021. | 2 | 2 | 2 | 2 | 2 | 1 | 1 | 1 | 2 | 2 | 17 |
| Utne et al., Midwifery, 2020. | 2 | 2 | 2 | 2 | 2 | 1 | 1 | 2 | 2 | 2 | 18 |
| Vang et al., Qualitative Health Research, 2018. | 2 | 2 | 2 | 2 | 2 | 0 | 2 | 0 | 0 | 2 | 14 |
| [Wah et al., Women and Birth, 2019.](https://www.sciencedirect.com/science/article/pii/S1871519217303244?via%3Dihub) | 2 | 2 | 2 | 2 | 2 | 1 | 2 | 2 | 1 | 2 | 18 |
| [Wilson et al., BMC Public Health, 2019.](https://www.ncbi.nlm.nih.gov/pmc/articles/PMC6434850/) | 2 | 2 | 2 | 2 | 2 | 2 | 2 | 2 | 2 | 0 | 18 |
| Results of updated search | | | | | | | | | | | |
| Faulks et al., WOMBI, 2024 | 2 | 2 | 2 | 2 | 2 | 2 | 2 | 2 | 2 | 2 | 20 |
| George et al., Midwifery, 2024. | 2 | 2 | 2 | 2 | 2 | 1 | 2 | 2 | 2 | 2 | 19 |
| Nechaeva et al., Global Health Action, 2024. | 2 | 2 | 2 | 2 | 2 | 0 | 2 | 2 | 2 | 2 | 18 |
| Parker et al., Qualitative Health Research, 2025. | 1 | 2 | 2 | 2 | 2 | 2 | 2 | 2 | 2 | 2 | 19 |
| Pierce et al., Front. Digit. Health., 2023. | 2 | 1 | 2 | 2 | 1 | 0 | 2 | 1 | 2 | 2 | 15 |
| Yuill et al., Br. J. Health Psychol., 2024. | 2 | 2 | 2 | 2 | 2 | 0 | 2 | 2 | 2 | 2 | 18 |

### Table S5: Mapping of sub-themes to Candidacy components

| **Candidacy component and contributing themes** | **Mapped sub-themes** | **N (%) mapped sub-themes** |
| --- | --- | --- |
| **Identification** |  |  |
| 1. Autonomy, dignity and personhood | 1.2 Acceptance of sub-par care | 3 () |
| 2. Informed choice and decision making | 2.1 Insufficient information |  |
|  | 2.3 Personalised counselling |  |
| **Navigation** |  |  |
| 2. Informed choice and decision making | 2.1 Insufficient information | 9 (26) |
|  | 2.2 Authoritative knowledge struggle |  |
|  | 2.3 Personalised counselling |  |
| 3. Trust in and relationship with HCP | 3.1 Stigma and mistrust |  |
|  | 3.2 Early initiation, relational care, and practical support |  |
| 4. Differences in healthcare systems and cultures | 4.1 Conceptualisation of pregnancy |  |
|  | 4.2 Lack of cultural competency |  |
|  | 4.3 Systems knowledge and social capital |  |
| 5. Systemic barriers | 5.1 Structural inadequacies |  |
| **Permeability of services** |  |  |
| 3. Trust in and relationship with HCP | 3.1 Stigma and mistrust | 6 (21) |
|  | 3.2 Early initiation, relational care, and practical support |  |
| 4. Differences in healthcare systems and cultures | 4.2 Lack of cultural competency |  |
|  | 4.3 Systems knowledge and social capital |  |
| 5. Systemic barriers | 5.1 Structural inadequacies |  |
|  | 5.2 Environmental factors |  |
| **Appearances at health services** |  |  |
| 1. Autonomy, dignity and personhood | 1.1 Not listened to | 5 (16) |
|  | 1.2 Wish to be seen as an individual |  |
| 4. Differences in healthcare systems and cultures | 4.1 Conceptualisation of pregnancy |  |
|  | 4.3 Systems knowledge and social capital |  |
| 5. Systemic barriers | 5.1 Structural inadequacies |  |
| **Adjudication** |  |  |
| 2. Informed choice and decision making | 2.1 Insufficient information | 5 (16) |
|  | 2.3 Personalised counselling |  |
| 3. Trust and relationship with HCP | 3.1 Stigma and mistrust |  |
|  | 3.2 Early initiation, relational care, and practical support |  |
| 5. Systemic barriers | 5.2 Environmental factors |  |
| **Offers and resistance** |  |  |
| 4. Differences in healthcare systems and cultures | 4.1 Conceptualisation of pregnancy | 2 (5) |
|  | 4.2 Lack of cultural competency |  |
| **Local production of candidacy** |  |  |
| 3. Trust and relationship with HCP | 3.2 Early initiation, relational care, and practical support | 3 (8) |
| 4. Differences in healthcare systems and cultures | 4.3 Systems knowledge and social capital |  |
| 5. Systemic barriers | 5.2 Environmental factors |  |

### Table S6: An extended Candidacy framework

| **Candidacy Framework factor** | **Definition** | **Individual-level** | **Health system-level** |
| --- | --- | --- | --- |
| **Existing factors** |  |  |  |
| Identification | Self-acknowledgement of necessity of medical attention for symptoms | ● | - |
| Navigation | Learning about and negotiating services | ● | ● |
| Permeability | Ease with which people can use services | - | ● |
| Appearance at health services | Individuals’ ability to articulate their need for care and assert their candidacy | ● | - |
| Adjudication | Health care-providers’ judgements dictating progression of individuals’ candidacy | - | ● |
| Offers & resistance | Declining to accept care offers, medications or referrals | ● | ● |
| Local production of candidacy | Local factors influencing candidacy, including availability of resources and long-term patient-provider relationships | ● | ● |
| **New factor proposed** |  |  |  |
| Intercultural dissonance | Additional barriers faced by those who are not native-born and experience a distinct difference in social norms and culture, medical and social knowledge and expectations, language, changing intergenerational relationships. | ● |  |
| Hostile bureaucracy | Discriminatory policies in the host-country, both in and outside of healthcare, that impose additional barriers such as restrictions on health coverage, welfare support, and right to rental properties; high visa application costs; and limits on qualifying employment. |  | ● |
